# First-in-human evaluation of memory-like NK cells with an IL-15 super-agonist and CTLA-4 blockade in advanced head and neck cancer

**DOI:** 10.1101/2024.09.16.24313758

**Authors:** Roman M. Shapiro, Michal Sheffer, Matthew A. Booker, Michael Y. Tolstorukov, Grace C. Birch, Moshe Sade-Feldman, Jacy Fang, Shuqiang Li, Wesley Lu, Michela Ansuinelli, Remy Dulery, Mubin Tarannum, Joanna Baginska, Nishant Dwivedi, Ashish Kothari, Livius Penter, Yasmin Z. Abdulhamid, Isabel E. Kaplan, Dinh Khanhlinh, Ravindra Uppaluri, Robert A. Redd, Sarah Nikiforow, John Koreth, Jerome Ritz, Catherine J. Wu, Robert J. Soiffer, Glenn J. Hanna, Rizwan Romee

**Affiliations:** Division of Transplantation and Cellular Therapies, Dana-Farber Cancer Institute, Harvard Medical School; Department of Informatics and Analytics, Dana-Farber Cancer Institute; Krantz Family Center for Cancer Research, Department of Medicine, Massachusetts General Hospital; Broad Institute of MIT and Harvard, Cambridge, MA 02142, USA; Translational Immunogenomics Lab, Dana-Farber Cancer Institute, Boston, MA, USA; Sorbonne University, Department of Clinical Hematology and Cellular Therapy, Saint-Antoine Hospital, Assistance Publique - Hôpitaux de Paris, Inserm UMRs 938, Centre de recherche Saint-Antoine, Paris, France; Center for Immuno-oncology, Dana Farber Cancer Institute, Harvard Medical School; Cell Transplant Therapy, CareDx Inc, Brisbane, CA, USA; Department of Surgery, Brigham and Women’s Hospital, Boston, MA, USA; Department of Data Science, Dana-Farber Cancer Institute

## Abstract

**C**ytokine ***i***nduced ***m***emory-***l***ike natural killer (CIML NK) cells combined with an IL-15 super-agonist (N-803) are a novel modality to treat relapsed/refractory head and neck cancer. We report data from a phase I trial of haploidentical CIML NK cells combined with N-803 with or without ipilimumab (IPI) in relapsed/refractory head and neck cancer patients after a median of 6 prior lines of therapy. The primary endpoint was safety, which was established, with dose-limiting toxicity in 1/10 patients. A promising though transient disease control rate of 70% correlated with donor NK cell expansion, the latter occurring irrespective of IPI. High-resolution immunophenotypic and transcriptional profiling characterized the NK cells and their interacting partners *in vivo*. IPI was associated with contraction of Treg:Tcon, rapid recovery of recipient CD8^+^ T cells, and accelerated rejection of donor NK cells. These results inform evaluation of CIML NK therapy for advanced malignancies, with considerations for combination with IPI.

**Significance:** CIML NK cells combined with N-803 and ipilimumab to treat head and neck cancer is safe, and associated with a more proliferative NK cell phenotype. However, the combination leads to reduced HLA mismatched NK cell persistence, resulting in an important limitation informing future NK cell combination therapies in clinical trials.

**Trial Registration:** NCT04290546

**Funding:** Gateway for Cancer Research, Ted and Eileen Pasquarello Research Fund, Goldfarb and Rudkin Family Fellowship, Dr. Antin’s Research Fund, Massey Family Fund. MS is funded by the Sidney Farber Scholar Program, Dana Farber, and RR is a recipient of the Career Development Award from the Leukemia and Lymphoma Society and is a member of the Parker Institute for Cancer Immunotherapy at Dana-Farber Cancer Institute.

## Introduction

Head and neck cancer represents the sixth most common malignancy worldwide, accounting for 3% of all annual new malignancy diagnoses and with an annual incidence that has been rising over the past decade.^1^ Relapsed/refractory head and neck cancer patients have a poor prognosis^2^ and represent a population in need of novel therapies. Building on the clinical benefit observed with PD-1-directed immunotherapy^3,4^, the adoptive transfer of natural killer (NK) cell-based therapy represents one such novel immunotherapeutic platform^5^, particularly as a high NK cell infiltrative signature has been associated with a favorable prognosis in head and neck cancer.^6^

Cytokine-induced memory-like (CIML) NK cells have increased capacity for *in vivo* expansion and persistence and demonstrate enhanced cytokine production and cytotoxicity to tumor targets.^7–9^ The clinical translation of CIML NK cell therapy in early phase trials has shown promising results in myeloid disease.^10–12^ The application of CIML NK cells for the treatment of relapsed/refractory head and neck cancer represents a natural evolution of this immunotherapy platform to a setting where it may offer therapeutic benefits when other options are limited.

IL-15 is a gamma-chain family cytokine that plays a key role in the differentiation and activation of NK cells.^13^ N-803 is an IL-15 superagonist (IL-15sa) that consists of an IL-15 mutein with enhanced activity, an IL-15R-α sushi domain mimicking physiologic trans-presentation, and an IgG1-Fc scaffold which substantially increases its half-life.^14^ This agent is associated with enhanced NK cell expansion and function *in vivo*, and has shown efficacy in prior trials leading to its approval in non-muscle invasive bladder cancer recently.^15–18^

In the current study, we sought to determine the safety of haploidentical donor-derived CIML NK cells in combination with N-803 with and without CTLA-4 blockade in a phase I trial. While IL-15 signaling does not expand Tregs, the recovery of these immunomodulatory cells following lymphodepletion and CIML NK cell infusion may potentially inhibit the activity of adoptively transferred donor NK cells. As Tregs are known to constitutively express high levels of CTLA-4,^19^ in the context of active CD16^+^ NK cells, these cells may be depleted by ipilimumab (IPI) through antibody-dependent cellular cytotoxicity (ADCC).^20^ We, therefore, hypothesized that enhanced ADCC function of the CIML NK cells in combination with IPI would mediate additional Treg depletion, leading to further enhanced CIML NK cell-mediated activity in the context of head and neck cancer. We observed that the addition of IPI was associated with contraction of Treg:Tcon in the peripheral blood lymphocyte compartment and promoted the expansion of mature and thus more cytotoxic NK cells early after adoptive transfer, but was also associated with more rapid NK cell rejection, thus acting as a “double-edged sword”. The safety and potential disease control seen in this study make a strong case for further evaluation of CIML NK cells plus IL-15sa. We posit that our findings will serve to guide future efforts aimed at combining NK cell-based approaches with other novel immunomodulatory agents.

## Results

### Adoptive transfer of haploidentical donor-derived CIML NK cells was safe and associated with disease control

This phase I clinical trial involved the infusion of haploidentical donor-derived CIML NK cells expanded and activated *in vivo* with N-803 following a course of lymphodepletion with fludarabine and cyclophosphamide. After completing the safety lead-in phase (n=6), the subsequent patients (n=4) also received a single dose of IPI (3mg/kg) on day −7 as a means to further enhance donor NK cell activity (hypothesized to occur via Treg depletion) (**Fig. 1a**). Altogether, eleven patients with head and neck cancer refractory to a median of 6 lines of prior therapy, including 8 patients with oropharyngeal primaries, were enrolled, and 10 patients received donor CIML NK cells (one patient died from disease progression before CIML NK infusion) (**Supplementary Figure 1**). The baseline patient and infusion product characteristics, as well as responses, are detailed in **Supplementary Table 1**. Overall, the treatment was well tolerated, meeting the primary safety endpoint of the trial. Among the first 3 patients in the cohort without IPI, one patient experienced septic shock secondary to pseudomonas infection leading to death and meeting criteria for a dose-limiting toxicity (DLT). An additional 3 patients were subsequently treated in the same cohort without IPI and no DLTs were observed. No DLTs were observed in the next cohort of patients treated with IPI. Among all the treated patients, lymphodepletion was associated with transient grade 3-4 pancytopenia, with peripheral blood counts recovering at a median of 13 days after CIML NK infusion. Five patients (50%) developed mild (grade 1-2) cytokine release syndrome (CRS) and responded to tocilizumab.

**Figure 1.**
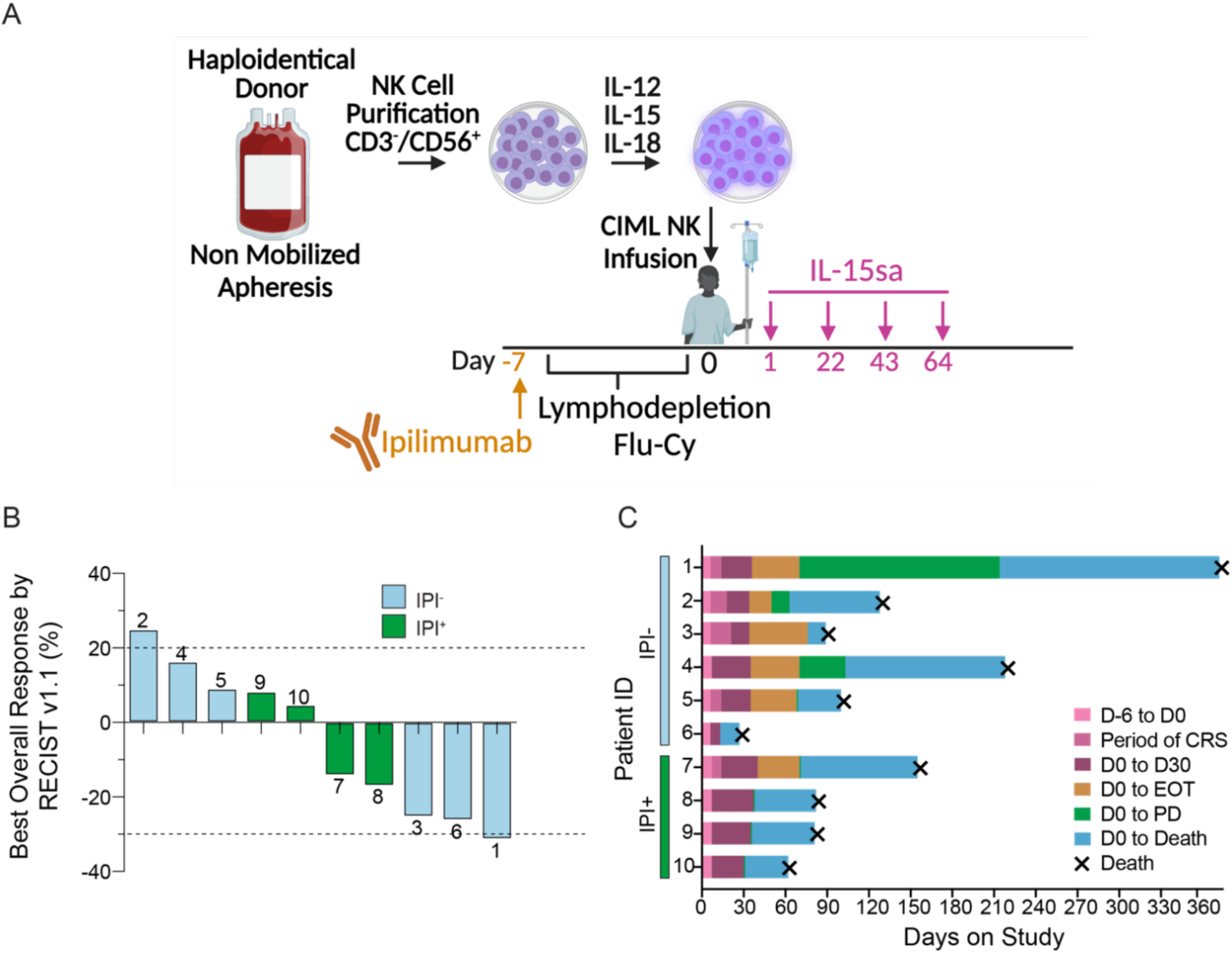
Clinical outcomes of patients with relapsed/refractory head and neck cancer treated with haploidentical donor-derived CIML NK cell therapy. A Clinical trial schema describing CIML NK infusion product generation and trial therapy. For each patient, a haploidentical donor underwent non-mobilized apheresis of peripheral blood followed by CD3 depletion and CD56 positive selection. The product was then incubated in a cocktail of IL-12, IL-15, and IL-18 for 12-16 hours to induce CIML differentiation and infused on day 0. The patients received lymphodepletion with fludarabine and cyclophosphamide, and IL-15 superagonist (IL-15sa) N-803 on day +1, with subsequent doses given every 3 weeks (up to 4 doses). After the first 6 treated patients were evaluated for safety, the subsequent patients also received a single dose of ipilimumab (IPI) 1mg/kg on day −7 prior to starting lymphodepletion (created using Biorender). B Waterfall plot showing tumor response by RECIST v1.1 criteria on day +30 following NK cell infusion. The bars in blue represent the first 6 evaluable patients on the trial who did not receive IPI while the green bars represent IPI treated patients. The number above each bar represents the patient ID on the trial. The dashed lines indicate size thresholds per RECIST v1.1 criteria. C Clinical course of all evaluable treated patients. Numbers on the vertical axis represent patient ID on the trial. The horizontal axis refers to days on study from the time of first therapy. D0: the day of CIML NK infusion, D30: day +30 response evaluation time point, CRS: cytokine release syndrome, EOT: end of treatment, PD: progressive disease.

Secondary endpoints on the trial included objective response rate (ORR), disease control rate (DCR), progression-free survival (PFS), and overall survival (OS). Ten patients were evaluable on day +30 following CIML NK infusion. Six patients (60%) had stable disease as the best response, and 1 (10%) patient attained a partial response by RECIST v1.1 criteria,^21^ with target tumor volume shrinking by 30% (**Fig. 1b & Supplementary Table 1**). This resulted in DCR 70% and ORR 10%. One patient had a malignant pleural effusion with head and neck cancer cells detectable at the time of initiation of trial therapy that demonstrated the predominance of NK cells (88% of the lymphocyte compartment) on day +14 following CIML NK infusion and had no detectable malignant cells in the pleural fluid on day +28 (Patient 3, **Supplementary Table 2**). Three patients (30%) had disease progression (Patients 2, 8, 10) as best response. At a median follow-up of 20.2 months, the median progression-free survival (PFS) was 3.0 months (95%CI 1-3.9), with the median overall survival (OS) 3.6 months (95%CI 1-4.7). There was no difference in PFS or OS between IPI treated and untreated patients (**Supplementary Figure 1**).

### A*doptive transfer of CIML NK cells followed by N-803 resulted in expansion and persistence of donor NK cells*

We performed an in-depth evaluation of peripheral blood mononuclear cells following donor CIML NK infusion using a combination of multimodal assays from longitudinally banked samples (**Fig. 2A**). Adoptively transferred CIML NK cells expanded (increased their proportion of the total lymphocyte compartment by a minimum of 2-fold relative to the baseline trial screening time point within 14 days of NK cell infusion) in 8 of 10 patients, constituting a median of 85% (range: 34-98%) of all lymphocytes on day +7 versus 14% (range: 2-29%) at the trial screening time point (**Fig. 2B & Supplementary Figure 2A**). Patients 2 and 8 did not have NK cell expansion at any evaluable time point, with the latter patient noted to have a donor-specific anti-HLA antibody (**Supplementary Table 3**). On day +14, the NK cells constituted a median of 70% (range: 1-92%) of the lymphocyte compartment in the patients with NK expansion (**Fig. 2B**). The median absolute NK cell counts at the time of screening (prior to CIML NK infusion), on day +7 and on day +14 were 77/uL, 157/uL, and 339/uL, respectively. NK cell expansion correlated with tumor volume reduction (**Supplementary Table 4**), with a higher fold expansion of the NK cell compartment (as absolute counts or percentage of lymphocytes) following infusion associated with a greater decline in tumor volume by RECIST v1.1 criteria (R = −0.71) (**Fig. 2C, Supplementary Figure 2B**). Donor NK cell expansion between days 7-14 following CIML NK infusion was confirmed by both flow cytometry by gating on mismatched HLAs; and by DNA-based chimerism assay employing next-generation sequencing (**Fig. 2D**). The NK cell compartment contracted to pre-infusion levels by day +28 in 8 of 10 patients, concurrent with a loss of donor chimerism, while 2 of 10 subjects displayed donor NK cell persistence beyond day +28 (**Fig. 2D**). There was a trend toward greater expression of IFNγ following *in vitro* cytokine stimulation by day +14 following donor CIML NK infusion, consistent with the memory-like phenotype of these cells (**Fig. 2E**).

**Figure 2.**
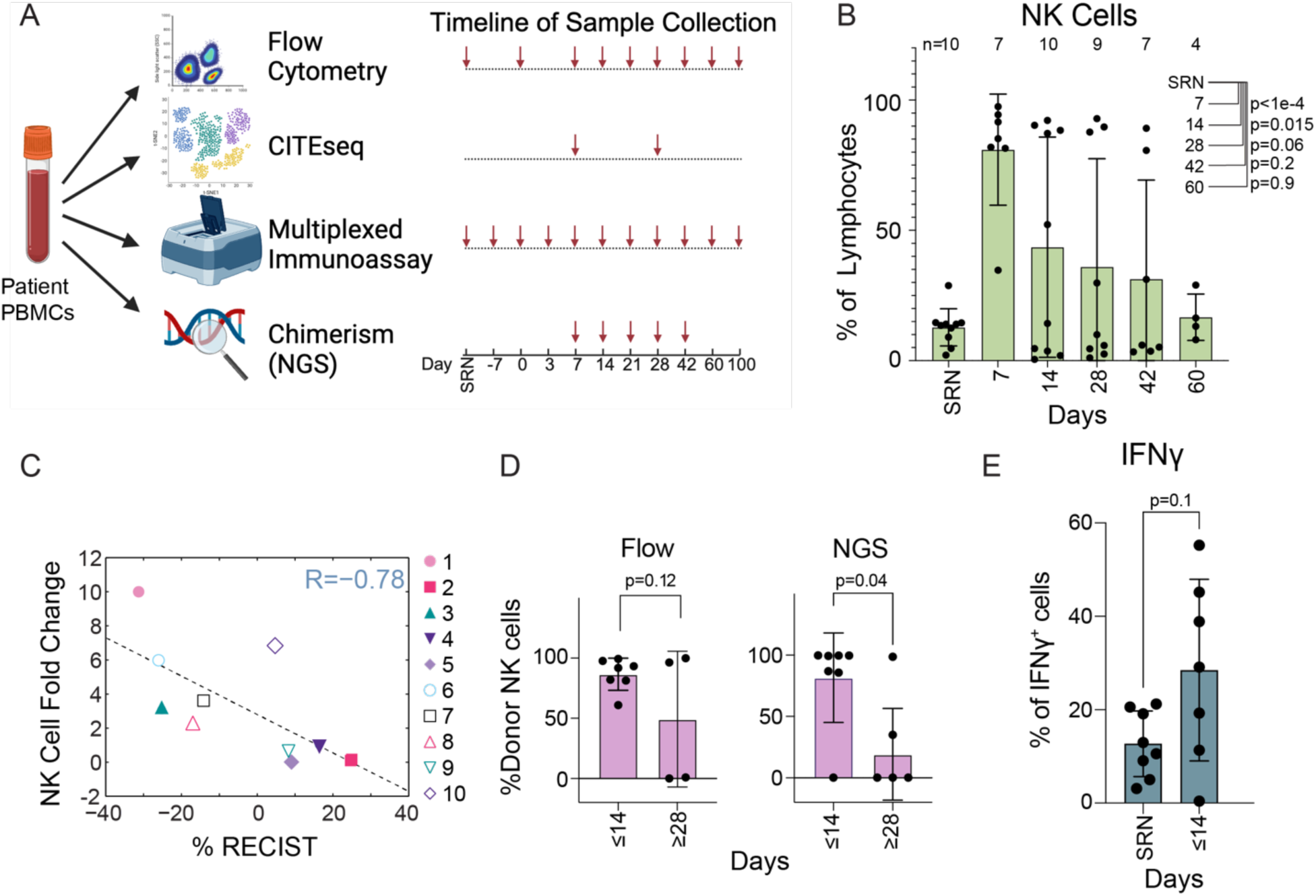
Expansion of the NK cells following donor cell product infusion. **A** Longitudinal evaluation of peripheral blood mononuclear cells (PBMCs) using flow cytometry with a customized panel, single-cell CITE-seq, a multiplexed immunoassay for secreted molecules, and next-generation sequencing of donor chimerism. The number of samples used for each assay at each time point is indicated (created using Biorender). **B** Size of the NK cell compartment as a proportion of total lymphocytes at the indicated time points after donor CIML NK infusion. Number of evaluable samples at each time point is shown above each bar. Adjusted p-values are determined using Dunnett’s multiple comparisons test. **C** Correlation between NK cell expansion and tumor responses by RECIST v1.1 criteria. NK cell fold change was calculated by taking the ratio of NK cells as a percentage of lymphocytes, in the peripheral blood on day 14 following CIML NK infusion to the percentage of NK at the time of screening. The patient ID is indicated at each point. **D** Donor cells as a proportion of all peripheral blood mononuclear cells at the indicated time points using two different assays. Flow cytometry-based evaluation (Flow) using anti-HLA antibodies targeting recipient-donor disparity in HLA. DNA-based chimerism using next-generation sequencing (NGS) performed by CareDx was also used to evaluate donor chimerism. ≤14 (n=7) refers to available samples collected between day +7 and day +14, ≥28 (n=4 for Flow, n=5 for NGS) refers to available samples collected between day +28 and day +60. **E** Percentage expression of interferon gamma (IFNγ) in flow cytometry based NK cell functional assays using PBMCs collected at the indicated time points following CIML NK cell infusion. p-value for all indicated comparisons were calculated using Mann-Whitney U. SRN: screening time point for the trial before NK cell infusion.

### NK cell expansion and persistence were characterized by early proliferative and later mature activated phenotypes

While NK cells were the dominant cell type in the lymphocyte compartment until day +14, CD8^+^ T cells reconstituted and replaced the NK cells as the dominant lymphocyte type by day +28 (**Fig. 3A**). We evaluated the immunophenotype of the NK cells in the peripheral blood over time following CIML NK infusion by flow cytometry, using a customized NK cell panel (**Supplementary Table 5**). Within the NK cell compartment, the majority were CD56^dim^ at all assessed time points, with a subset expressing NKG2A, a subset expressing KIR, and another expressing CD57 (**Fig. 3B**).^22,23^ While NKG2A^+^ and KIR^+^ cells constituted a stable proportion of CD56^dim^ NK cells in the peripheral blood after CIML NK infusion, the proportion of terminally differentiated CD57^+^ NK cells declined over time (**Fig. 3B**).

**Figure 3.**
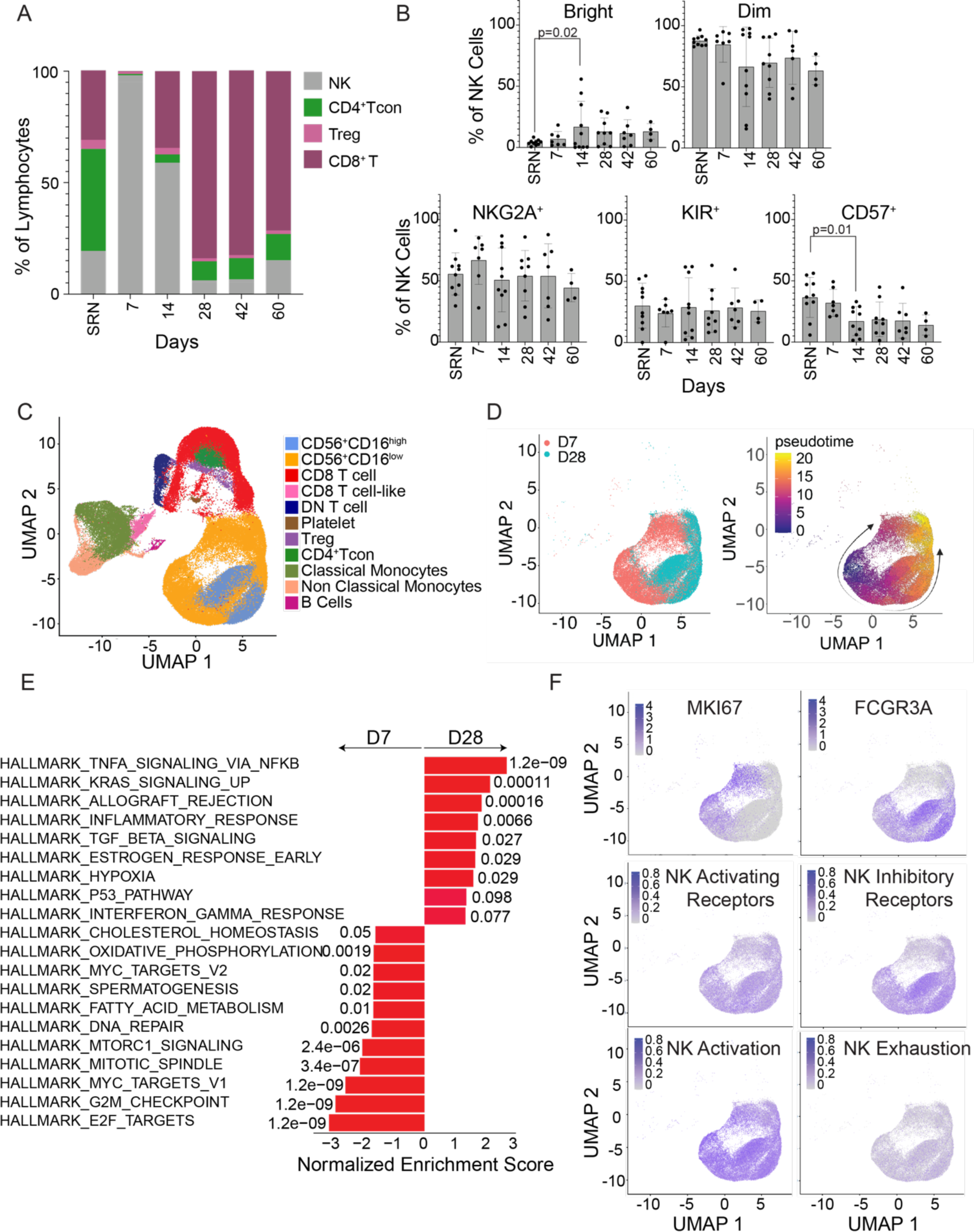
NK cell populations exhibit markers of proliferation and activation following CIML NK infusion. **A** Distribution of NK cells and T cells in the peripheral blood as a proportion of total lymphocytes assessed with flow cytometry. Data presented is a percentage of total lymphocytes. Gating strategies are indicated in **Supplementary Figure 17**. **B** Distribution of the CD56^dim^ (Dim) NK cell populations and various subsets (NKG2A^+^, KIR^+^, CD57^+^) as well as the CD56^bright^ (Bright) NK cell population assessed with flow cytometry, mean and SD. Adjusted p-values for each time point comparison with SRN are determined using Dunnett’s multiple comparisons test and detailed in **Supplementary Table 13**, significant p-values are indicated. SRN: screening time point **C** UMAP of NK cell clusters defined using single-cell CITE sequencing data, combining all patient samples (n=4) and all time points following CIML NK infusion (D7: day +7, D28: day +28). **D** Pseudotime plots demonstrating the differentiation trajectories of expanding NK cells. The plot on the left shows the same UMAP clustered by time. **E** Gene set enrichment analysis of pathways in NK cell clusters comparing days +7 (D7) and +28 (D28), with the earlier time point showing enrichment of proliferative gene sets while the latter time point showing enrichment of pathways associated with NK cell activation. **F** Expression of markers of proliferation (*MKI67*), *FCGR3A* (gene for CD16) and gene lists of NK activating receptors, inhibitory receptors, activation, and exhaustion. Key genes comprising these lists are described in the supplemental appendix (**Supplementary Table 7**). The intensity of expression is represented by the color in the legend within each plot.

To evaluate the proteomic and transcriptional profiles of the adoptively transferred CIML NK cells and study their potential interactions with other immune cells *in vivo* at single-cell resolution, we performed single cell CITE-seq, which combines single cell RNA sequencing with a custom oligonucleotide tagged antibody panel^24^ (**Fig. 3C & Supplementary Table 6**). We found that NK cells underwent a transcriptional and immunophenotypic shift following their *in vivo* expansion (**Fig. 3D**). On day +7, gene set enrichment analysis showed evidence of NK cell proliferation (MYC_TARGETS_V1) (**Fig. 3E**) as well as a signature of NK activation (i.e. *JUN, JUNB, FOS, FOSB, GZMB*, and *PRF1;* **Fig. 3F & Supplementary Table 7**). Furthermore, gene expression representative of CD56^bright^ (*CD44*), adaptive (*CD3ε, IL32*), and mature CD56^dim^ (*GZMB, NCR3*) NK cells were present (**Supplementary Figure 3**).^23^ In contrast, the patterns of gene expression on day +28 were consistent with the presence of predominantly mature CD56^dim^ NK cells (**Supplementary Figure 3**), with the expression of transcripts associated with NK cell activation^25^ (**Fig. 3E**) and NK cell inhibitory receptors (i.e. KIR3DL1, NKG2A, and *KLRB1;* **Fig. 3F, Supplementary Figure 4 & Supplementary Table 7**).

The *MKI67* gene was among the most differentially expressed in NK cells on day +7 compared to day +28. Other genes most highly expressed on day+7 included vimentin (*VIM*), stathmin 1 (*STMN1*), and tubulin (*TUBA1B*) -- all involved in cytoskeletal regulation and mitotic cell cycle (**Supplementary Figure 5**). There was a minimal expression of an NK exhaustion signature, characterized by genes such as *HAVCR2 (Tim-3) and PDCD1*. NK cells at all times expressed activating receptors, including *KLRK1*, NKp30, and NKp46. Concurrently, the largest concentration of known NK cell ligands for the expressed receptors was observed on the monocyte clusters (**Supplementary Figure 4**). The TGFβ signaling pathway was upregulated in NK cells on day +28, compared to day +7 (**Fig. 3E**), characterized by the expression of *JUNB*, *HIPK2*, and *ID2* (**Supplementary Figure 6**). Other key genes more highly expressed in NK cells on day +28 included the NK inhibitory receptor *KLRB1*, as well as *ZFP36, TXNIP, and SPON2* involved in cytokine responses and regulation as well as inflammatory cellular stress (**Supplementary Figure 5**).^26–28^

Pseudotime analysis suggested two distinct differentiation trajectories of the infused NK cells (**Fig. 3D**): on day +7, one of these trajectories led to a CD16^lo^ NK cell population characterized by an upregulation of cell cycle genes (**Supplementary Figure 7**) and proliferation (**Fig. 3F**), while the other trajectory led to a CD16^hi^ population of predominantly mature CD56^dim^ NK cells expressing markers of NK activation concurrent with a gene expression signature comprised of NK inhibitory receptors (**Fig. 3F & Supplementary Figure 7**). Upregulation of the chemokine genes *CCL3* and *CCL4*, and a chemotaxis gene signature characterized by the integrin receptor subunit genes *ITGA1* and *ITGA2* were also noted as the NK cell differentiated following infusion (**Supplementary Figure 8**).^29^

### Distinct patterns of secreted molecules were associated with NK cell expansion and concurrent changes in the tumor burden

We sought to evaluate the secretory molecule milieu to determine whether these molecules supported NK cell expansion and persistence and correlated with tumor burden reduction. Using a panel of 92 markers (**Supplementary Table 8**) applied to longitudinally collected plasma samples, we found distinct patterns of secreted molecules following NK cell infusion (**Fig. 4A**). Endogenous soluble IL-15 levels (without measurement of N-803) were increased early after CIML NK infusion, concurrent with the period following lymphodepletion (**Fig. 1A & Fig. 4B**).^30^ IL-6 levels were also concurrently increased, coinciding with the clinical development of CRS (though mild in all cases) in treated patients (**Fig. 1C & Fig. 4B**). IL-18 levels, while initially depressed after infusion, recovered toward day +14. IL-12 followed a similar pattern but with a distinct delay, resulting in a relative depletion of this cytokine from the plasma during the period of donor NK cell expansion and persistence. Increased plasma concentrations of CXCL10, CCL3, and CCL4 also coincided with the period of NK cell expansion, while that of CD8A was relatively reduced. The concentration of IFNγ and Granzyme B in the plasma of all treated patients was increased by day +7 and gradually decreased by day +28, concordant with the pattern of donor NK cell chimerism (**Fig. 4B**).

**Figure 4.**
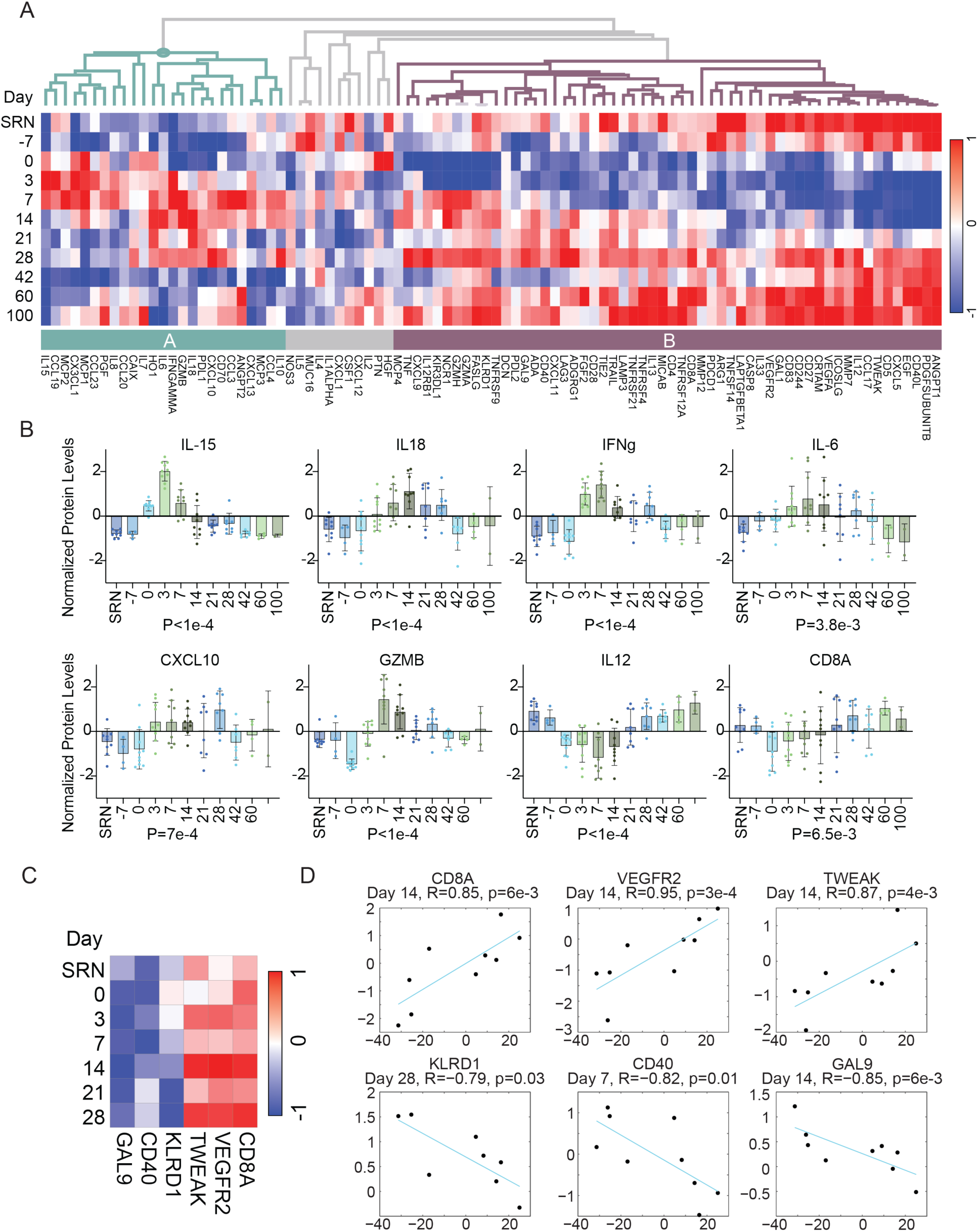
Distinct patterns of secreted molecules following CIML NK infusion support NK cell growth and correlated with tumor burden. **A** The O-link panel (92 markers) was used to evaluate the secretion of cytokines into the serum of evaluated patients at various time points as indicated on the y-axis. Patterns of secreted molecules clustered into those that were upregulated at the time of maximal NK expansion (A, turquoise bar) and those that were downregulated at the time of maximal NK expansion (B, purple bar). **B** The concentration of specific secreted molecules in plasma over time is shown, mean and SD. A comparison between maximal secreted molecule concentration and screening time point (SRN) is shown (Mann-Whitney U), with p-values indicated for each molecule. **C** Differentially secreted molecules positively correlated with tumor burden increase (red) and tumor burden decrease (blue). **D** Correlation between secretion of molecules at the indicated time points and tumor responses by RECIST v1.1 criteria. Relative expression levels are shown on the y-axis and tumor volume change on the x-axis.

We next correlated the secreted molecule patterns with tumor volume change by RECIST v1.1 criteria. There was no correlation between the plasma concentrations of IL-12, IL-15, IL-18, GZMB, or IFNγ with the change in tumor burden following CIML NK infusion. We found that increased plasma concentrations of the NK receptor KLRD1 and the molecules GAL9 and CD40 correlated with the reduction in tumor volume, while increased plasma concentrations of the molecules CD8A, VEGFR2, and TWEAK (TNFSF12) were associated with tumor volume increase (**Fig. 4C**). The strongest positive correlations of secreted molecules with tumor volume increase occurred on day +14, while those correlating negatively with tumor volume occurred at various time points following NK cell infusion (**Fig. 4D**).

### The addition of ipilimumab was associated with increased NK cell proliferation, but an earlier contraction of the NK cell compartment following donor CIML NK infusion

To investigate the impact of IPI on immune reconstitution, we evaluated NK and T cell populations longitudinally and compared IPI non-treated (IPI^-^) and treated (IPI^+^) patients in this trial. NK cell expansion as a proportion of total lymphocytes by day +14 was not different between IPI^-^ and IPI^+^ groups (**Fig. 5A**), but treatment with IPI was associated with a trend toward more rapid absolute NK cell expansion (p=0.06) (**Supplementary Figure 9**), and a relative increase in mature CD16^+^CD57^+^KIR^+^ NK cells by day +7 (**Supplementary Figure 10 & Supplementary Table 9**). However, at the same time, IPI was associated with an earlier contraction of the NK cell compartment, with more rapid recovery of CD8^+^ T cells by day +28 in these patients (**Fig. 5A**). Furthermore, a comparison of peripheral blood mononuclear cell populations on days +7 and +28 based on single cell CITE-seq showed that the IPI treatment associated with persistence and earlier recovery of T cell and monocyte populations (**Fig. 5B**). Cell populations segregated as distinct populations on days +7 and +28 based on exposure to IPI treatment (by UMAP), suggesting that peripheral blood CD16^hi^ NK cells were depleted in the IPI^+^ patients (**Fig. 5C**).

**Figure 5.**
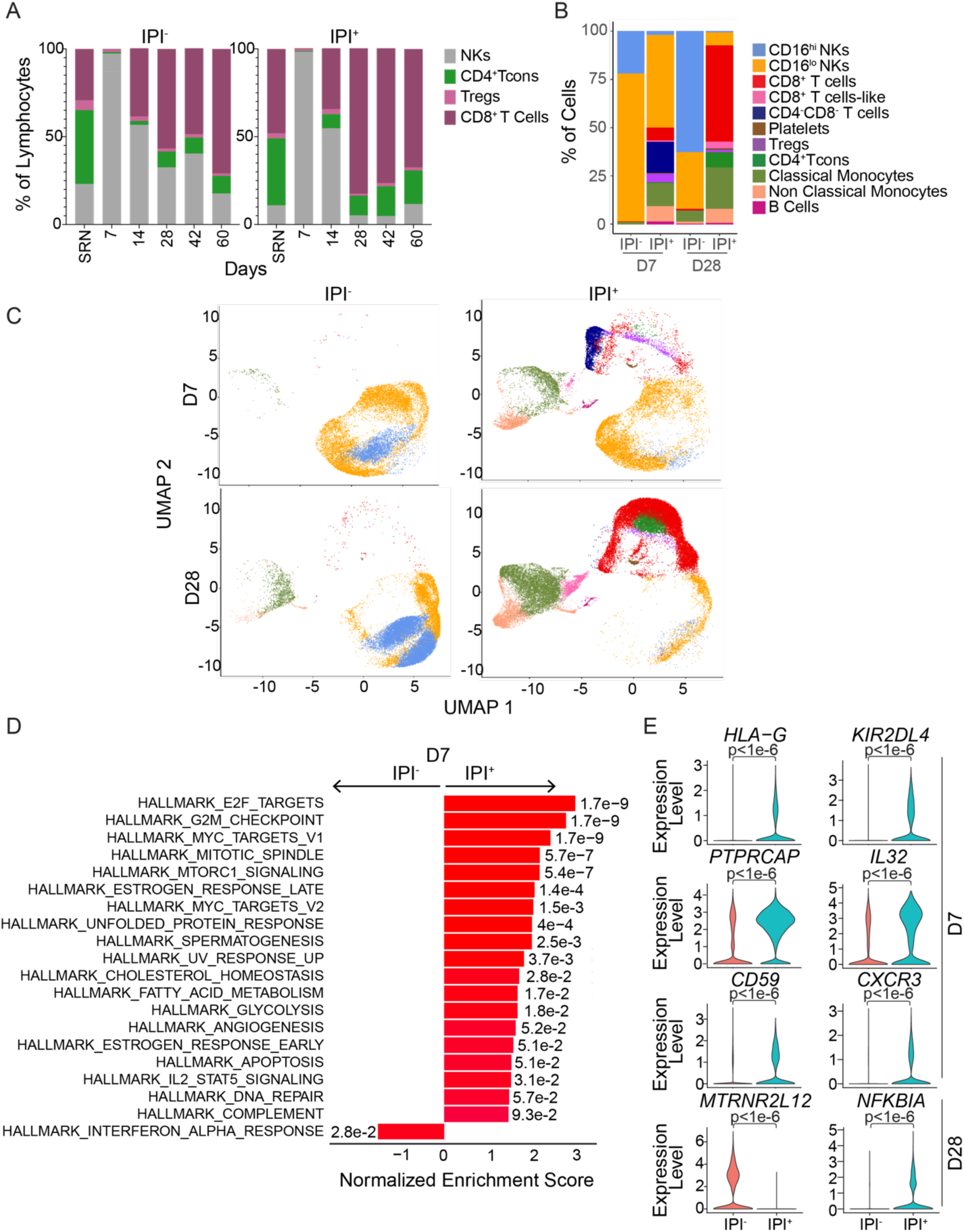
**Ipilimumab is associated with increased early NK cell proliferation but more rapid contraction of the NK cell compartment**. **A** Distribution of NK cells and T cells in the peripheral blood as a proportion of total lymphocytes, stratified by IPI treatment, as evaluated with flow cytometry. The data presented is a percentage of total lymphocytes. **B** Distribution of peripheral blood mononuclear cells as measured with single cell CITE-seq on day +7 (D7) and day +28 (D28), comparing IPI treated (IPI^+^, n=2) and IPI untreated (IPI^-^, n=2) patients. **C** UMAP of NK cell clusters defined with single cell CITE-seq, separated by time points assessed and IPI treatment. The color scheme corresponds to the same populations as shown in B. **D** Gene set enrichment analysis of pathways in NK cell clusters comparing IPI^+^ (n=2) and IPI^-^ (n=2) patients on day +7 (D7). **E** Most differentially expressed genes in NK cells comparing IPI treated and untreated conditions on day +7 (D7) and day +28 (D28). Adjusted p-values were determined with Mann-Whitney U test.

Regarding the impact of IPI treatment on NK functional states, no clear association was observed however the use of IPI (**Supplementary Figure 11**) was associated with the expression of transcripts of receptors that may regulate NK cell effector function, including *KIR3DL1*, *KLRD1, KLRC1, HAVCR2, and ITGAL/LFA-1* (**Supplementary Figure 12 and Supplementary Tables 10 & 11**). IPI was also associated with the upregulation of several key genes on day +7 that included proliferative gene sets in NK cell populations (**Fig. 5D**), NK activation coreceptor CD59, KIR2DL4, and its ligand HLA-G which is associated with a proinflammatory NK cell phenotype,^33^ cytotoxicity promoting cytokine IL-32,^34^ and *CXCR3* a key NK cell trafficking gene^35^ (**Fig. 5E**) concurrent with increased secretion of its cytokine ligand CXCL10 in patient plasma (**Fig. 4B**). The expression of mitochondrial lncRNA (*MTRNR2L12*), which is associated with reduced apoptosis,^36^ was noted in the persisting NK cells in the non-IPI (IPI^-^) patients on day +28, while *NFKIB* was upregulated in the IPI^+^ patients (**Fig. 5E**).

### Ipilimumab treatment was associated with a decreased Treg:Tcon ratio and increased reconstitution of activated CD8^+^ T cells

To determine how adoptively transferred NK cells may interact with recipient T cells in mediating their effects and how IPI may potentially affect this interaction, we evaluated T cell reconstitution in all trial patients. Both CD4^+^ Tcon and CD8^+^ T cells were depleted on day +7 in all patients, with relative persistence of Treg (**Fig. 6A**). CD8^+^ T cells exhibited recovery on day +14 and became the dominant T cell population by day +28, with most having the central memory phenotype (**Supplementary Figure 13**). A comparison of PBMC populations on days +7 and +28 with CITE-seq was consistent with the flow cytometry data, further showing distinct T cell clusters between the two time points. At the earlier time point, T cell clusters were predominantly double negative (DN) T cells and Tregs, while at the later time point, they were predominantly CD8^+^ and CD4^+^ Tcons (**Supplementary Figure 13 & Fig. 3C**). Mirroring the findings in NK cells, the day +7 time point was associated with the upregulation of proliferative pathways in T cells by GSEA while the day +28 time point was associated with an upregulation of inflammatory pathways, suggesting a state of T cell activation (**Fig. 6B**).^37^

**Figure 6.**
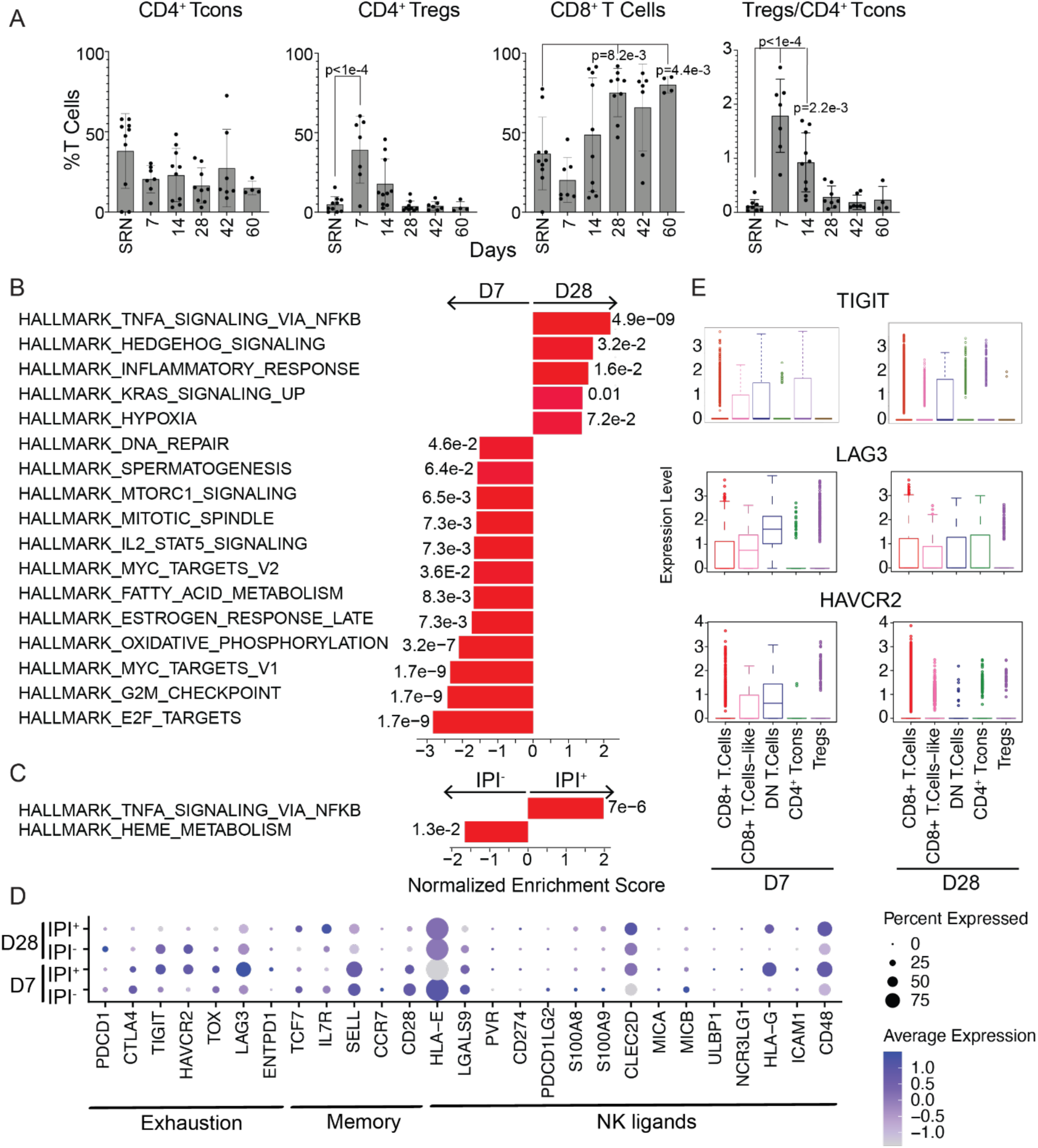
T cell reconstitution following CIML NK infusion. **A** Distribution of the T cell populations assessed by flow cytometry, mean, and SD. Adjusted p-values for each time point comparison with SRN (trial screening time point) are determined using Dunnett’s multiple comparisons test and detailed in **Supplementary Table 13**, significant p-values are indicated. **B** Gene set enrichment analysis of pathways in T cell clusters comparing days +7 (D7) and +28 (D28), showing enrichment of pathways of activation at the latter time point. **C** Gene set enrichment analysis of pathways in T cell clusters comparing IPI untreated (IPI^-^) and treated (IPI^+^) conditions. **D** Dot plot showing genes defining cell signatures of exhaustion and memory in T cell subpopulations, as well as NK ligands, separated by time and IPI treatment. Mann-Whitney U tests were performed per group of genes, adjusted p-values for each gene are detailed in **Supplementary Table 14**. **E** Expression of exhaustion markers in T cell populations on days +7 (D7) and +28 (D28). Boxplots show median and quartiles. Mann-Whitney U test. P-values were calculated per cell type and are detailed in **Supplementary Table 15**.

Since IPI was added to potentially deplete CTLA4^+^ Tregs in the context of donor memory-like NK cell infusion, we compared Treg reconstitution in the IPI^-^ and IPI^+^ contexts. We found that the IPI^+^ context had a relative reduction in Treg:Tcon on day +14 following CIML NK infusion (**Supplementary Figure 13**). On gene expression analysis, the TNFa signaling via NFkB pathway, which plays a role in enhancing costimulatory signaling to the T cells,^37^ was particularly associated with the IPI^+^ context (**Fig. 6C**). While there was a trend for greater expression of markers of T cell exhaustion on day +7 in the IPI^+^ context, there was no difference in the expression of markers of T cell memory between IPI^+^ and IPI^-^ patients (**Fig. 6D**). Exhaustion markers such as *TIGIT, LAG3* and *HAVCR2* were most notable in the DN T cell population. While some CD8^+^ and CD4^+^ Tcon expressed *LAG3* on day +28, most of these cells did not express exhaustion markers (**Fig. 6E**).

### Use of Ipilimumab was associated with MHC-I signaling interactions between NK and T cells

To explore whether the reconstituting T cell populations on both day +7 and day +28 exhibited evidence of crosstalk with the NK cells, we evaluated the expression of known NK ligands in these cells using single cell CITE-seq. At both time points, T cell populations expressed ligands for NK cells, including the activating ligand CD48 and lower levels of the inhibitory ligand HLA-E particularly in the IPI^+^ context (**Fig. 6D**). To further explore the potential effects of donor NK cell populations on reconstituting immune effector cells in the presence of N-803 and IPI, we conducted an interaction analysis using CellChat.^38^ In the absence of IPI, the CD16^lo^ NK cells, which constituted the majority of NK cells on day +7, had relatively few predicted cell-cell interactions with T cells in contrast to the CD16^hi^ NK cells (**Fig. 7A**). However, in the presence of IPI, far more predicted cell-cell interactions were detected between CD16^lo^ and CD16^hi^ NK cells and other immune effector cells (**Fig. 7A**). Prominent among these interactions was that of the NK inhibitory receptor *KLRB1* (gene for CD161), which was among the most highly differentially expressed genes in NK cells on day +28 (**Supplementary Figure 5**), and its ligand *CLEC2D* (gene for LLT1) on CD4^+^ Tcons (**Supplementary Figure 14**). Other potential interactions between T cells and NK cells were in the CCL axis (**Supplementary Figure 15**), with CCL3 and CCL4 gene expression being predominantly in NK cells (**Supplementary Figure 16**). The analysis also predicted cell-cell communication via the MHC-I signaling axis in the context of IPI treatment, which occurred between CD16^hi^ NK cells and CD4^+^ Tcons on day +7 (**Fig. 7B**) but was predominantly driven by the interaction between CD8^+^ T cells and CD16^hi^ NK cells on day +28 (**Fig. 7C**). The latter interaction was largely absent in the IPI^-^ patients, instead being largely driven by CD4^+^ Tcons. As the MHC-I signaling axis curated by the CellChat database mainly reflects interactions between the CD8 receptor and HLA ligands,^38^ in the HLA mismatched context this would suggest CD8^+^ T cell-mediated rejection.

**Figure 7.**
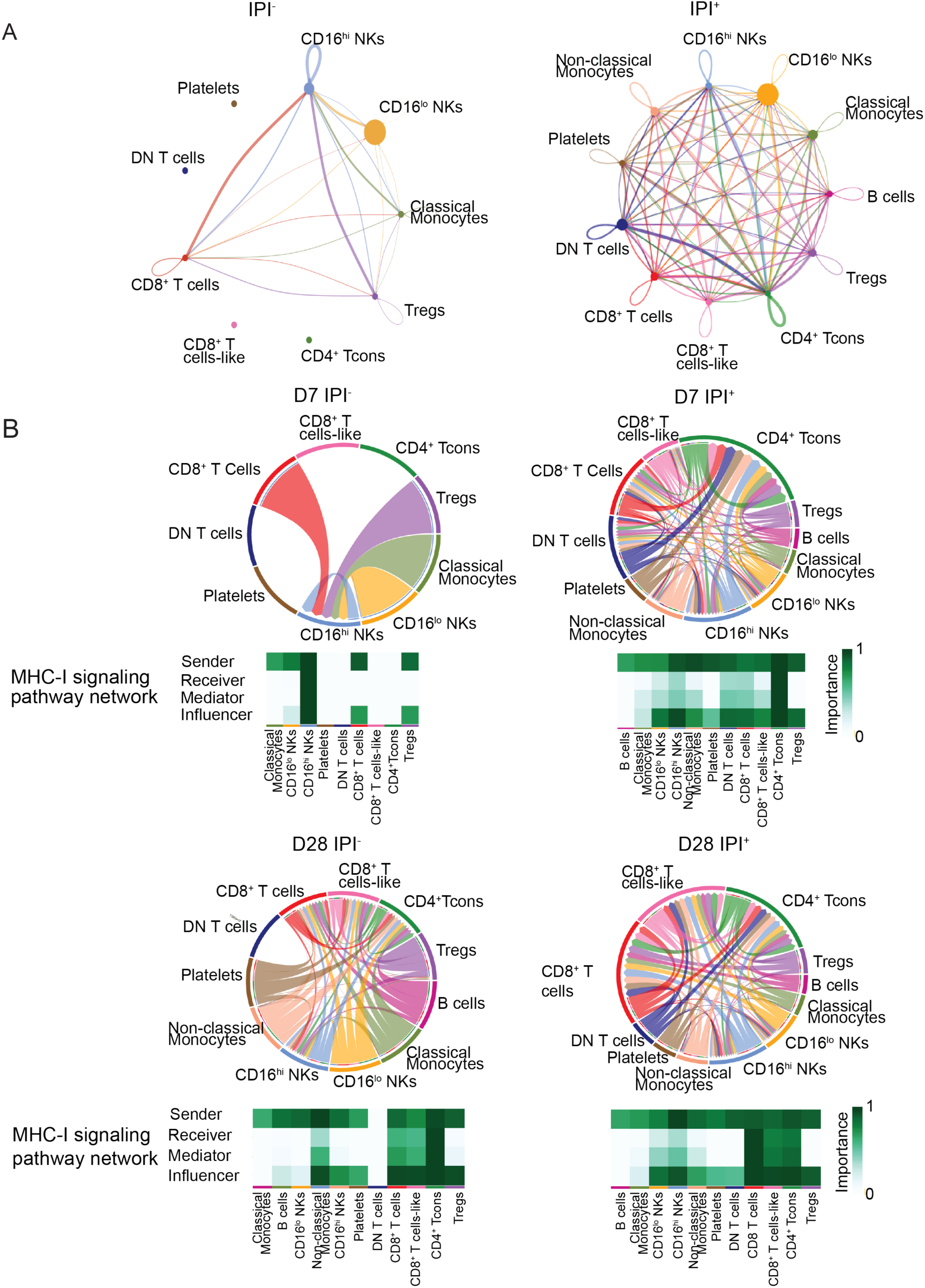
Cell-cell interaction analysis between CIML NK cells and T cells. **A** Cell-cell interaction analysis using CellChat applied to the CITE-seq data on day +7, showing the interaction maps in IPI^-^ and IPI^+^ patients. **B** Circos plots showing predicted cell-cell interactions via the MHC-I signaling pathway on day +7 (D7) in IPI^-^ and IPI^+^ patients. Predicted senders and receivers of signals in the MHC-I pathway as predicted with CellChat are indicated beneath each circos plot. **C** Circos plots showing predicted cell-cell interactions via the MHC-I signaling pathway on day +28 (D28) in IPI^-^ and IPI^+^ patients.

## Discussion

Herein, we demonstrate that the combination of donor CIML NK infusion in combination with N-803 with or without IPI in patients with advanced relapsed/refractory head and neck cancer is safe and associated with a promising though transient disease control. The disease control correlated with the NK cell expansion and an in-depth multi-omic analysis including CITE-seq allowed us to evaluate key phenotypic and gene expression changes in both the adoptively transferred NK cells and the recipient immune cell compartment. The addition of IPI was associated with decreased Treg:Tcon, increased activated CD8^+^ T cells, and a more rapid drop in the NK cells suggestive of enhanced rejection. We predicted novel immune cell-cell interactions which were further modulated by the addition of IPI. Overall, our findings not only support further evaluation of this innovative immunotherapy but also highlight its broad impact on multiple components of the immune cell compartment.

The addition of IPI in this trial stemmed from the hypothesis that it may induce antibody dependent cellular cytotoxicity (ADCC) targeting the CTLA4^+^ Tregs.^20^ While IPI treatment was associated with a reduced Treg:Tcon ratio and a more proliferative and activated NK cell phenotype, it also correlated with a larger recipient CD8^+^ T cell compartment by day +28 coinciding with a more rapid contraction of the peripheral blood donor NK cell compartment. While the donor NK cells persisted for a shorter period in the context of IPI, they did exhibit strikingly more predicted cell-cell interactions with other cells in the peripheral blood than in the IPI untreated patients. NK cells were the predicted receivers of immunomodulatory *CLEC*^40^ signaling via *KLRB1* in the context of IPI, with the senders of the signal being primarily CD4^+^ Tcons. Additionally, CD8^+^ T cells and CD4^+^ Tcons were also the major receivers of signaling in the CCL axis, which in the context of increased secretion of CCL3 and CCL4 in the plasma during the period of NK cell expansion and gene expression of these chemokines in NK cells suggested that the latter were the senders of at least some of this signaling.

It is worth noting that CD16^hi^ NK cells were the predominant interacting partners with other immune effector cells based on CellChat analysis, both with and without IPI. Coupled with the observation that persistence of this subgroup of NK cells on FlowSOM analysis of flow cytometry data was associated with more tumor burden reduction as measured with RECIST v1.1 criteria, the enhanced cross-talk between these NK cells and other immune effector cells may play an important role in NK cell therapy. The interplay between the IL-15sa and IPI in the cross-talk between NK cells, other immune effector cells and tumor requires further study.

This study has important limitations. First, while we observe a correlation between donor NK cell expansion and tumor burden reduction, the number of treated patients is small. Furthermore, even though NK cells constituted the majority of the lymphocyte compartment in 7/10 patients, there was some heterogeneity in the absolute NK cell numbers in the peripheral blood in spite of all patients receiving similar NK cell product doses. In addition, due to the clinical status of the relapsed/refractory patients enrolled on the trial, biopsy of tumor following CIML NK infusion was not feasible, leading to a paucity of tumor tissue samples. Although these limitations do not allow for simple generalizations of all the findings, the deep immunophenotypic and transcriptional analysis of longitudinal samples generated important hypotheses with potential for future clinical translation. The transient disease control observed in the study has prompted further amendments to the protocol, including the removal of lymphodepleting chemotherapy, extending the use of N-803 beyond day +100, addition of the epidermal growth factor receptor inhibitor cetuximab, use of an off-the-shelf NK cell product that allows for repeated infusions (Clinicaltrials.gov ID: NCT06239220), and consideration of using an autologous CIML NK cells to prevent their rejection. Ongoing work aims to validate and better understand the mechanisms driving inferred interactions between donor NK cells and recipient immune effector cells to further optimize the efficacy of the memory-like NK cell platform.

In summary, we demonstrate that the haploidentical donor CIML NK cells in combination with N-803 with or without IPI treatment is safe and associated with promising though transient disease control in patients with relapsed/refractory head and neck cancer. While IPI was associated with more proliferative and activated NK cells, it was also associated with shorter donor NK cell persistence likely driven by CD8^+^ T cell-mediated allogeneic rejection.

## Methods

### Trial participants

This phase 1 single-center (Dana-Farber Cancer Institute), open-label, clinical trial enrolled patients with recurrent or metastatic (R/M) head and neck cancer (either head and neck squamous cell carcinoma [HNSCC] or salivary gland cancer) regardless of human papillomavirus (HPV) status who had received prior platinum and immunotherapy (anti-PD-1) treatments in the R/M setting (for the HNSCC patients, specifically); with no restrictions of number of lines of prior therapy. Patients 18 years of age or older with any smoking history were eligible if their ECOG performance status was 2 or less and they had measurable disease (RECIST v1.1). The trial was approved by the DF/HCC institutional review board (IRB) (19-505), conducted in accordance with the Declaration of Helsinki and Good Clinical Practice Guidelines, and registered nationally (NCT04290546).

### Study design

Patients in cohort 1 received lymphodepleting chemotherapy with fludarabine (25 mg/m^2^/day) for 5 days starting on day −6 and cyclophosphamide (50 mg/kg/day) for 2 days starting on day −5 prior to haploidentical CIML NK cell infusion on day 0 (5-10 x 10^6^ viable cells/kg=dose level 0) followed by N-803 (IL-15 superagonist [IL-15sa], 15 mcg/kg subcutaneously) starting on day +1 every 21-days for 4-doses. In cohort 2, patients received the same regimen with a dose of lead-in ipilimumab (3 mg/kg IV) on day −7. Cohort 1 treated 3 patients at dose level 0 (adhering to a 3+3 dose de-escalation design); <2 DLTs triggered an additional 3 patients (n=6). Cohort 2 treated an additional 4 patients. The primary study objectives were to establish safety and tolerability and to estimate the maximum tolerated dose (MTD) of CIML NK cells. Secondary objectives were to characterize the objective response rate (RECIST v1.1) to therapy at day +30 (4 weeks) and day +84 (12 weeks) following CIML NK cell infusion, to estimate progression-free survival (PFS) and overall survival (OS), and to determine the phenotypic expansion and function of adoptively transferred NK cells with correlation to clinical benefit and survival. Data as of December 17, 2023, were analyzed. Deidentified clinical data that can be shared in accordance with institutional policy is available upon request from Dr. Glenn Hanna.

### Patient and healthy donor samples

Patients provided written informed consent prior to participation under an IRB-approved protocol at the Dana-Farber Cancer Institute where all protocol procedures were performed and data were collected. Correlative samples included peripheral blood collected at screening for the trial, on day −7 (before starting lymphodepletion), day 0, day +7, day +21–28, day +42, day +60, day +100, and relapse time points after infusion. Samples acquired from patients for the purpose of all correlative studies cannot be obtained again from the same patients. Healthy donor PBMCs (peripheral blood mononuclear cells) were isolated by Ficoll centrifugation, and NK cells were purified using EasySep (STEMCELL Technologies, 17955). All PBMCs were cryopreserved according to a standard internal laboratory protocol at the time of collection from patients. Exclusion of any collected samples from planned correlative studies was only done if the cell viability was poor or cell quantity was insufficient. Due to the clinical status of the heavily pretreated patients enrolled on this trial, collection of tumor biopsy samples for research purposes following CIML NK cell infusion was not feasible.

### Flow cytometric analysis

A custom NK and T cell panel of antibodies was used (**Supplementary Table 4**). All cell staining for flow cytometry was performed as previously described^10^, and data were acquired on a BD LSR Fortessa flow cytometer and analyzed using FlowJo (Tree Star) software. Gating strategies for NK cells and T cells are shown in representative plots (**Supplementary Figure 17**). The absolute lymphocyte count was measured using a clinical assay. For any lymphocyte population not measurable directly with the clinical assay, the absolute count of this population was determined by multiplying its percentage (percent of total lymphocytes) as measured with flow cytometry by the total lymphocyte count measured at the same time point using the clinical assay. For one patient we used day 21 sample instead of day 28 which was not available.

### Olink cytokine assays

Serum from the patients were analysed for 96 protein biomarkers using the Olink Target 96 Immuno-Oncology panel (**Table S8**). Olink proteomics is based on a proximity extension assay, where oligonucleotide-labeled antibody probe pairs are allowed to bind to their respective targets in the sample in 96-well plate format. The quality control was performed using Olink® NPX Signature software, and additional QC was performed by removing seven poorly detected proteins from further analyses. Data are presented as log2-transformed units NPX. Higher NPX values correspond to a higher protein expression. Spearman correlation was used to detect cytokines associated with response and cytokines were selected based on adjusted p<0.1.

### Functional and flow-based cytotoxicity assays

Frozen PBMCs were thawed and cultured overnight in RP10 medium (RPMI 1640 supplemented with 10% FBS, 1× penicillin/streptomycin, 2mM L-glutamine, and 7.5 mmol HEPES) with 1ng/mL IL-15. K562 cells were cultured in RP-10 medium and labeled with 5uM of CellTrace Violet (Thermo Fisher Scientific) in PBS for 20 min at 37°C and washed with RP10. To measure HLA donor chimerism, intracellular IFNγ and degranulation, cells were stained as follows: PBMCs, PBMCs with cytokines (IL-12 10ng/ml, IL-18 50ng/ml, IL-15 50ng/ml), or PBMCs with K562 cells at 10:1 E:T ratios, were co-cultured for 1h, followed by the addition of 0.2uL BD GolgiPlug™ (BD Biosciences), 0.13uL BD GolgiStop™ (BD Biosciences) and 1uL of APC-CD107a (Biolegend). After an additional 5h of co-culture, cells were stained for extracellular proteins with CD56, CD3, CD16, HLA antibodies (**Supplementary Tables 5,12**) and violet live-dead stain (thermos fisher, 1:1000). Intracellular staining for IFNγ was performed using BD Cytofix/Cytoperm (BD Biosciences). Cells were acquired using BD LSR Fortessa and analyzed using FlowJo (Tree Star). Data is presented selectively for donor NK cells, based on the staining of HLA.

### Donor Chimerism

The assessment for the persistence of the donor CIML NK cell therapy product in patient samples was performed using the AlloCell test (CareDx). NK cells were isolated from PBMCs using negative bead selection for CD56 (STEMCELLS technologies). Cell pellets were shipped to CareDx for AlloCell testing. Briefly, genomic DNA was extracted and measured by Qubit (Qubit™ Flex Fluorometer, Thermo Fisher Scientific) for quality control. A minimum of 8ng gDNA was used for targeted amplification of hundreds of single nucleotide polymorphisms (SNPs) in a single multiplex reaction on Mastercycler® Nexus Thermal Cycler (Eppendorf). Next Generation Sequencing (NGS) was performed on NextSeq 550 (Illumina). Percent cell product measurements were established by analyzing the relative contributions of SNP alleles with CareDx’s proprietary AlloCell data analysis pipeline incorporating open and custom bioinformatics tools. The results were reported as % Cell product relative to the patient genome.

The assessment for the persistence of the donor CIML NK cell therapy product in patient samples was also performed using flow cytometry evaluating HLA (**Supplementary Table 12**). Cells were stained for either donor or recipient as part of the functional assays described above.

### Single-cell RNA-seq and CITE-seq sequencing and analysis

Cryopreserved PBMC samples from patients were thawed and diluted in RP10 media. Samples with low viability rates were enriched for live cells using dead cell removal (Annexin V) kit (STEMCELLS technologies). Cells were stained with CITE-seq antibodies (TotalSeq™-C Human Universal Cocktail, V1.0, BioLegend) per manufacturer’s instruction, using 1:3 or 1:4 dilution of the antibody pool, supplemented with 2ul of each custom antibody. Briefly, 0.5-1 million cells were resuspended in 50uL cell staining buffer (PBS with 0.04% ultrapure BSA, #AM2616), 5uL of Human TruStain FcX Fc Blocking reagent (BioLegend) was added and the mixture was incubated for 10 minutes at 4°C. Following this, the CITE-seq antibody cocktail, adjusted to 50ul in staining buffer, was added and incubated for 30 minutes at 4°C. The mixture was then washed 3 times with 1 mL the staining buffer and resuspended in 1000 cells/uL. Single-cell libraries were prepared using the 10x Chromium Next GEM Single Cell 5’ v2 Kit (10XGenomics), according to the manufacturer’s instructions. The single-cell RNA-seq and CITE-seq libraries were sequenced by NovaSeq S4 flowcell (Illumina). Single-cell RNA-seq and Cite-seq data were processed, aligned to hg38, and aggregated using CellRanger (version 7.1.0).^50^ Cite-Seq and expression data were analyzed using Seurat (version 5.0.0).^51^ Data from individual samples was analyzed separately prior to combining the data from multiple samples. The outlier cells with a very low number of gene features (<200) or low total UMI (<500), or high number of gene features (>7000) were removed. Cells with a high mitochondrial ratio (>20%) from each data set were also removed. Subsequently, samples were combined based on the identified anchors for the following integrated analysis. Principal component analysis (PCA) was performed and the first 10 principal components (PCs) were used to cluster cells using the Louvain algorithm in Seurat. These PCs were then used to run UMAP clustering. Well-defined marker genes for each cluster were used to identify potential cell populations, such as T cells (CD2, CD3D, CD3E, CD3G), Treg (RTKN2, FOXP3, CD4, IL2RA, TIGIT, CTLA4, FCRL3, LAIR2, IKZF2), B cells (IGLL5, MZB1, JCHAIN, DERL3, SDC1, MS4A1, BANK1, PAX5, CD79A), CD14^+^ monocytes (S100A9, CTSS, S100A8, LYZ, VCAN, S100A12, IL18, CD14, G0S2, FCN1), CD16^+^ monocytes (CDKN1C, FCGR3A, PTPRC, LST1, IER5, MS4A7, RHOC, IFITM3, AIF1, HES4) and NK cells (NCAM1, KLRD1, KLRF1, NKG7, GNLY), platelets (PPBP, PF4, NRGN, GNG11, CAVIN2, TUBB1, CLU, HIST1H2AC, RGS18, GP9). Differential gene expression between scRNA-seq cell population was performed using a pseudo-bulk approach with patients as replicates and DESeq2 (version 1.34.0).^52^ Interaction data was analyzed using CellChat (version 1.6.1).^38^ Pseudotime analysis was performed using Monocle3 (version 1. 3.4).^53^ For gene sets representing specific cellular functions or pathways, we performed functional enrichment analysis using the Hallmark gene set collection.^54^

### Statistical Analyses

All flow cytometry data were tested for normal distribution (Shapiro-Wilk test), and if the data were not normally distributed, the appropriate non-parametric tests were used (GraphPad Prism v5.0). Differential expression of single-cell RNA sequencing data was performed using the Mann-Whitney U test as part of the Seurat package.^51^ All statistical comparisons are indicated in the figure legends. All comparisons used a two-sided alpha of 0.05 for significance testing. All measurements were taken from distinct samples. All data for correlative studies and analysis is available upon request from the corresponding author.

### Study Approval

This study was reviewed and approved by the institutional review board of the Dana-Farber Cancer Institute, Boston, MA, USA (Clinicaltrials.gov: NCT04290546). The study was performed in compliance with the provisions of the Declaration of Helsinki and Good Clinical Practice guidelines. Written informed consent was obtained from participants before inclusion in the study.

## Data Availability

All data produced in the present study are available upon reasonable request to the authors

## Author Contributions

RMS and MS contributed equally to the study. GH and RR designed the clinical study protocol. RMS, MS, and RR designed the correlative studies. SN prepared CIML NK products under good manufacturing practice (GMP) guidelines and was responsible for quality control of the products. GB, YZA, IEK, DK, MuT processed and prepared patient samples for correlative studies, and performed experiments. MS performed the NK cell functional assays. RMS, MS, ND, and AK planned and conducted NGS-based chimerism using patient samples. MA performed flow cytometry and RMS, MS, RD and MA analyzed the flow cytometry data. LP, RMS, and MS planned the single cell CITE-seq experiments. MS, MSF, JF, SL, and WL performed single-cell CITE-seq, and RMS, MS, MAB, and MYT performed data analyses. JB performed the Olink cytokine panel. GH recruited clinical trial participants. RMS and MS analyzed all the data and designed the figures. RMS, MS, GH, and RR interpreted the data and wrote the first draft of the manuscript. CJW, JK, RJS, RU and JR reviewed the manuscript and provided critical feedback on the results of the correlative studies. All authors revised the manuscript critically and approved the final version.

## Acknowledgements

We would like to thank M. Shipp (DFCI) for discussion and guidance, and D. Hearsey (DFCI) for assistance in processing and banking clinical samples. We are grateful to Miltenyi Biotech for providing materials for cell processing and product preparation. We are grateful to ImmunityBio for providing N-803, and to BMS for providing IPI. We thank Allyson Moulton, BS, Evgeniya Vaskova, PhD Natali Gulbahce, PhD, Marica Grskovic, PhD, and the R&D lab at Cell Transplant Therapy, CareDx, Inc., for their assistance with planning, developing, performing and analyzing the results of the NGS-based chimerism assay used in this manuscript. We are grateful for the patient volunteers who gave their time and provided the samples for all correlative studies. We are also grateful to the cellular therapy and research coordinators at the Dana-Farber Cancer Institute.

