## Supplementary Data for "First-in-human evaluation of memory-like NK cells with an IL-15 super-agonist and CTLA-4 blockade in advanced head and neck cancer"

**Supplemental Data**

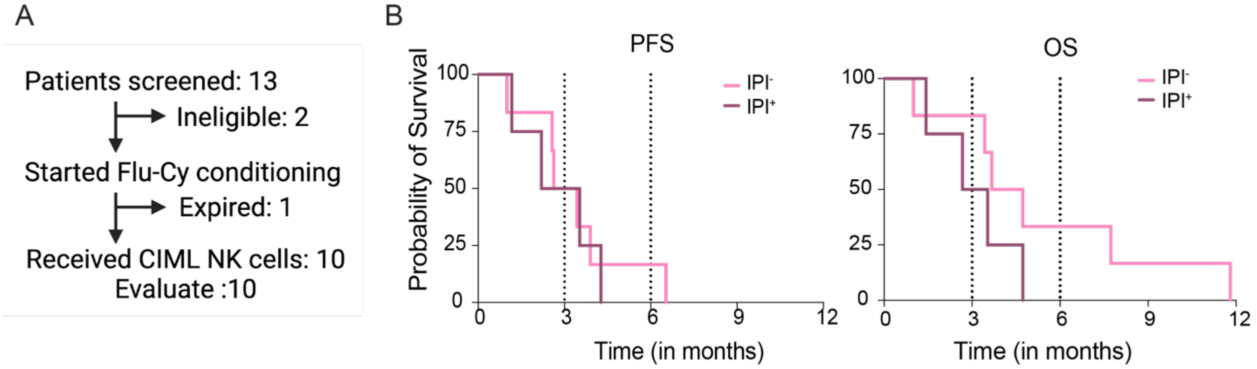


**Figure S1. Phase I clinical trial outcomes. A** CONSORT diagram for patient screening and enrolment. **B** Kaplan-Meier curves for patients treated in the study, showing progression-free survival (PFS) and overall survival (OS) for patients in the IPI untreated (IPI$^-$, n=6) and IPI treated (IPI$^+$, n=4) groups.

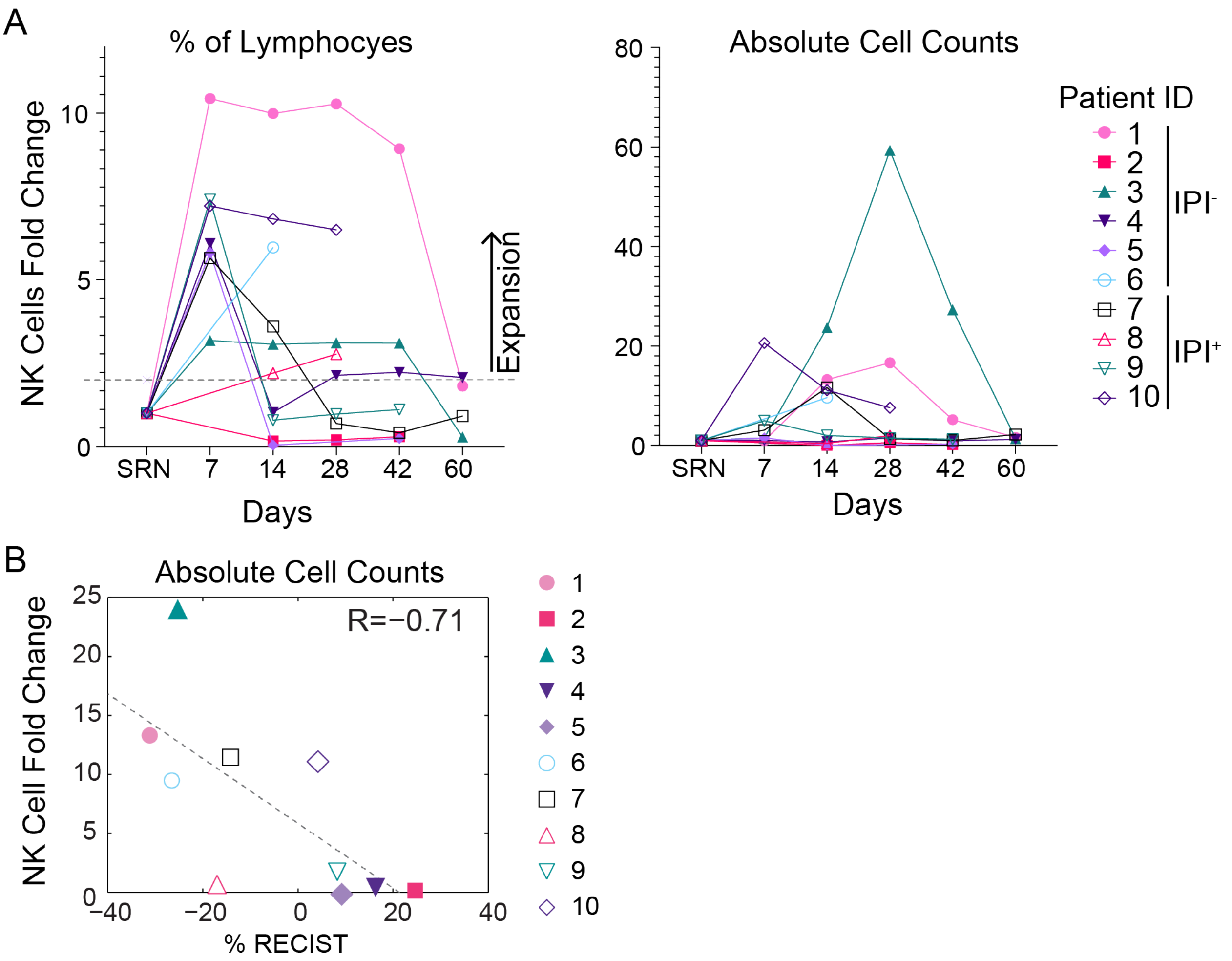


**Figure S2. NK fold expansion in patients receiving CIML NK cells with N-803. A** Fold expansion was determined using the percentage of NK cells out of total lymphocytes (left) or absolute counts of NK cells (right), calculated as the ratio between the different time points and the SRN (trial screening) time point prior to CIML NK infusion. Individual patient NK fold expansion is indicated using colors corresponding to patient ID on the trial. Dashed line indicates 2-fold as the threshold above which NK cells were considered expanded. **B** Correlation between NK cell expansion and tumor responses by RECIST v1.1 criteria. NK cell fold change was calculated by taking the ratio of absolute NK cell numbers in the peripheral blood on day +14 following CIML NK infusion to the absolute NK cell numbers in the peripheral blood at the time of screening. The patient ID is indicated at each point.

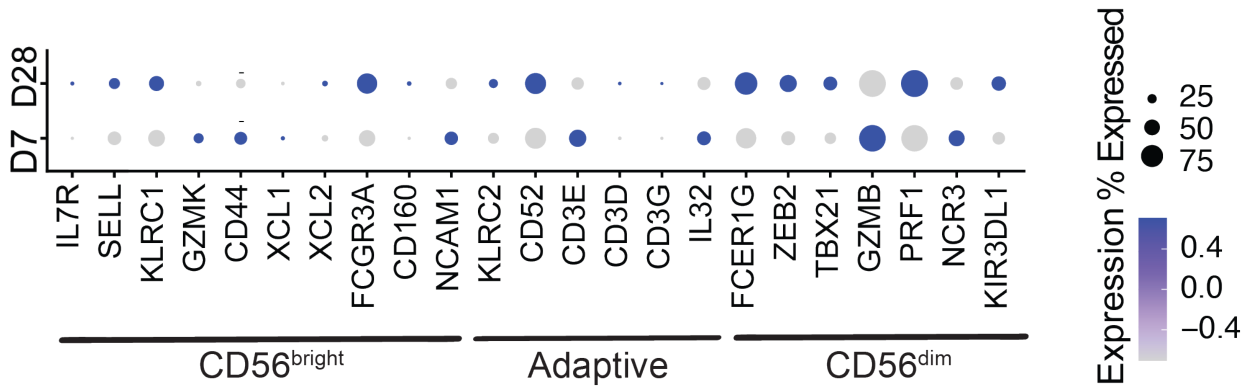


**Figure S3. Dot plot of gene expression of NK cell markers over time.** Gene expression analysis with single cell CITEseq applied to peripheral blood NK cells in patients (n=4) groups on day +7 (D7) and day +28 (D28) following donor CIML NK infusion. The size of the dots reflects the number of cells expressing a gene, and the color reflects the degree of expression. Genes are grouped based on expression in known types of NK cells. P-values were calculated using Mann-Whitney U test are detailed in Supplementary Table 14.

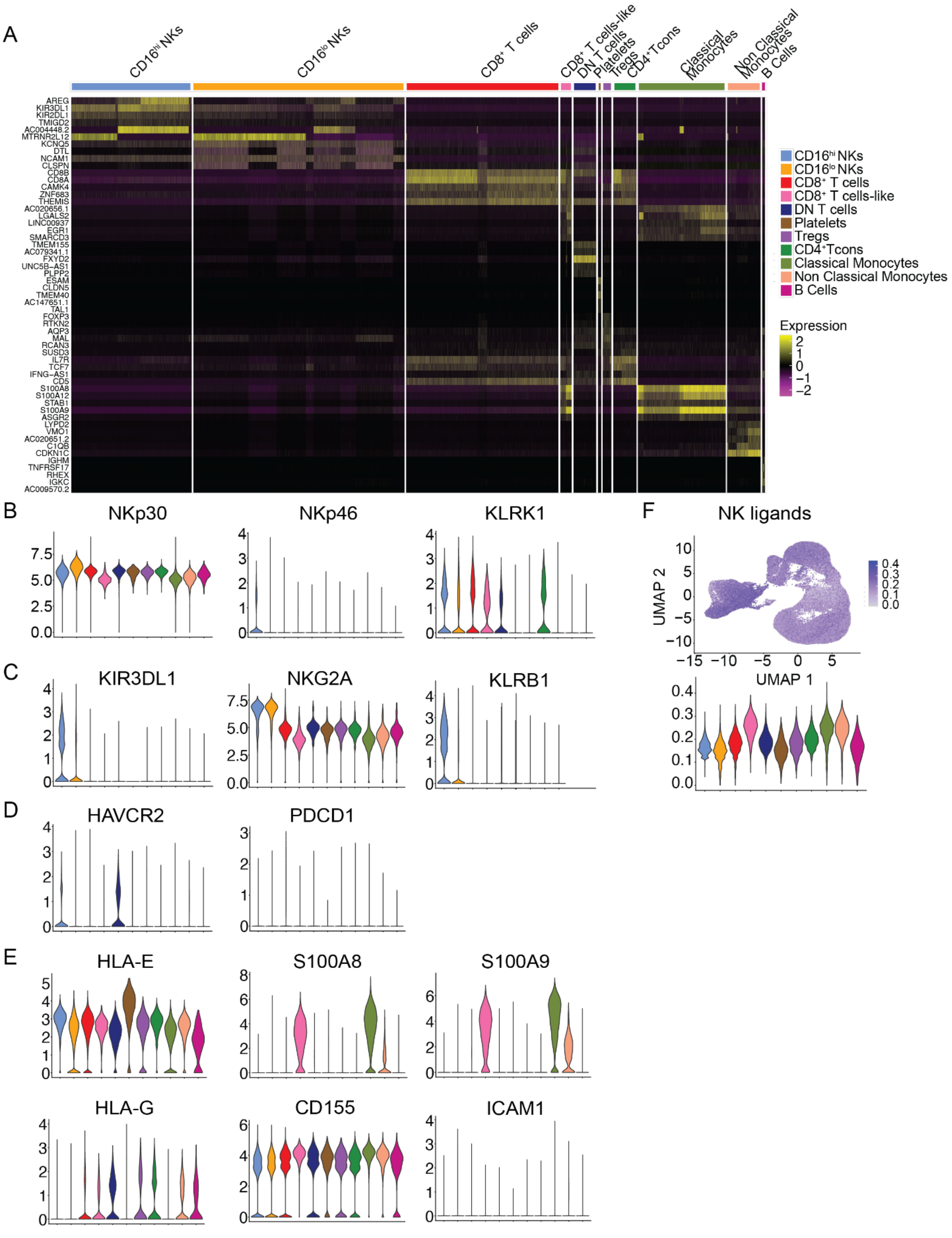
A
CD16hi NKs
CD16lo NKs
CD8+ T cells
CD8+ T cells-like
DN T cells
Platelets
Tregs
CD4+Tcons
Classical Monocytes
Non Classical Monocytes
B Cells
AREG
KIR3DL1
KIR2DL1
TMIGD2
AC004448.2
MTRNR2L12
KCNQ5
DTL
NCAM1
CLSPN
CD8B
CD8A
CAMK4
ZNF683
THEMIS
AC020656.1
LGALS2
LINC00937
EGR1
SMARCD3
TMEM155
AC079341.1
FXYD2
UNC5B-AS1
PLPP2
ESAM
CLDN5
TMEM40
AC147651.1
TAL1
FOXP3
RTKN2
AQP3
MAL
RCAN3
SUSD3
IL7R
TCF7
IFNG-AS1
CD5
S100A8
S100A12
STAB1
S100A9
ASGR2
LYPD2
VMO1
AC020651.2
C1QB
CDKN1C
IGHM
TNFRSF17
RHEX
IGKC
AC009570.2
Expression
2
1
0
−1
−2
B
NKp30
NKp46
KLRK1
C
KIR3DL1
NKG2A
KLRB1
D
HAVCR2
PDCD1
E
HLA-E
S100A8
S100A9
HLA-G
CD155
ICAM1
F
NK ligands
UMAP 2
UMAP 1

**Figure S4. Characteristics of NK cells in the peripheral blood following CIML NK infusion. A** Heatmap of key genes characterizing each of the major cell clusters. **B** Violin plots of select NK activating receptors in all clusters. **C** Violin plots of select NK inhibitory receptors in all clusters. **D** Violin plots of select exhaustion markers in all clusters. **E** Violin plots of select NK ligands in all clusters. **F** Expression of a summative NK ligand signature in all cells. The greatest concentration of NK ligands was noted in monocytes clusters.

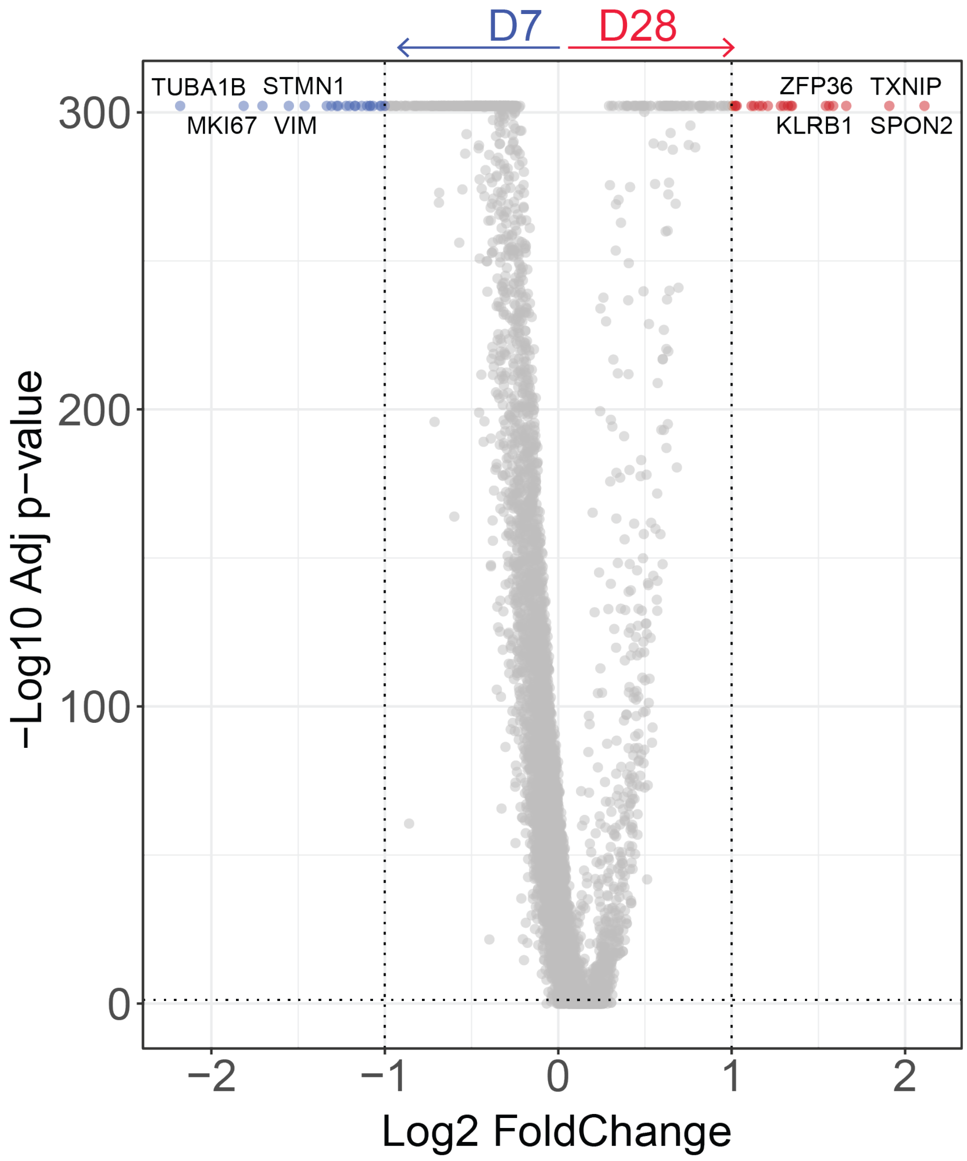


**Figure S5. Differential gene expression in NK cells clusters over time.** Genes expressed more highly in the NK cells on day 7 (D7) are shown on the left (blue), and genes expressed more highly in the NK cells on day +28 (D28) are shown on the right (red).

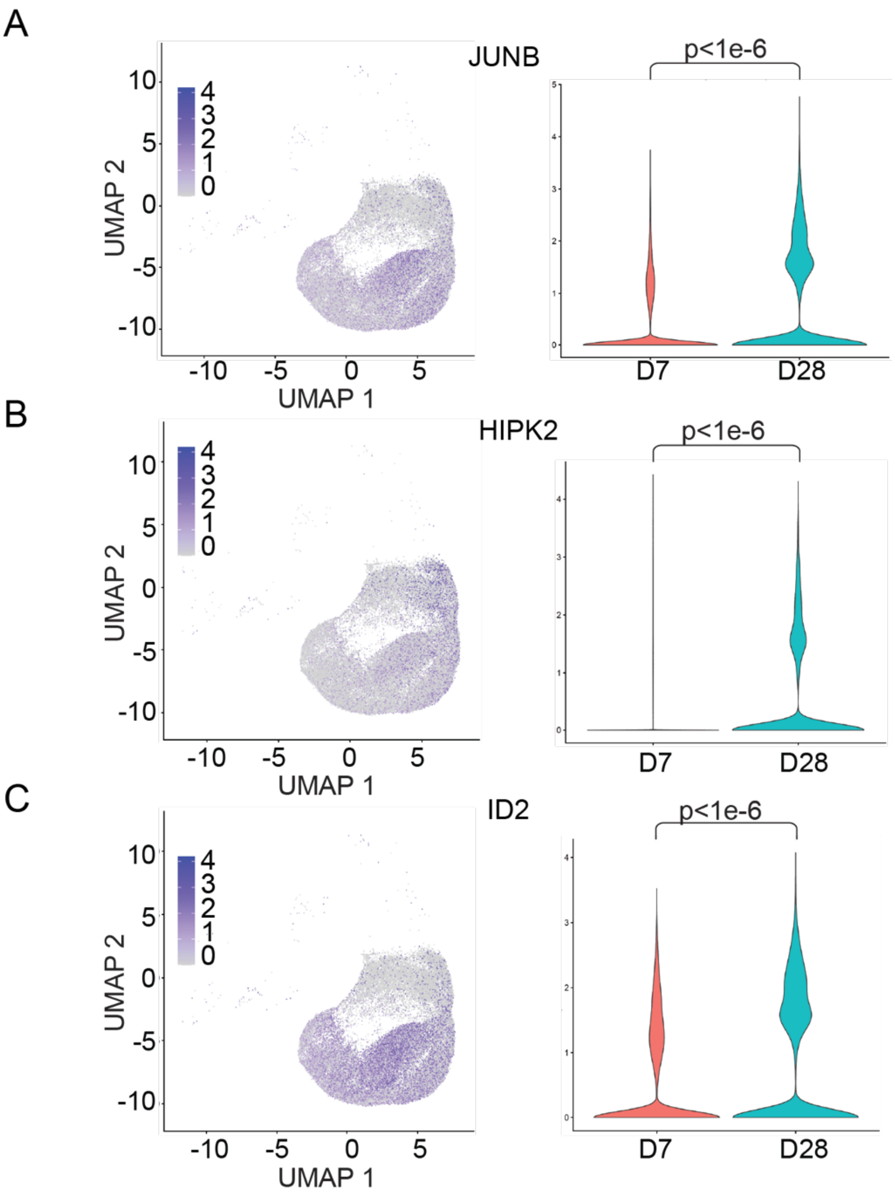


**Figure S6. Expression of TGFβ pathway genes in NK cells, with the most differentially expressed genes between day +7 and day +28 shown. A** Expression of JUNB in NK cell clusters on UMAP and in violin plots on day +7 (D7, red) and day +28 (D28, turquoise). **B** Expression of HIPK2 in NK cell clusters on UMAP and in violin plots on day +7 (D7, red) and day +28 (D28, turquoise). **C** Expression of ID2 in NK cell clusters on UMAP and in violin plots on day +7 (D7, red) and day +28 (D28, turquoise). Mann Whitney U test, adjusted p-values.

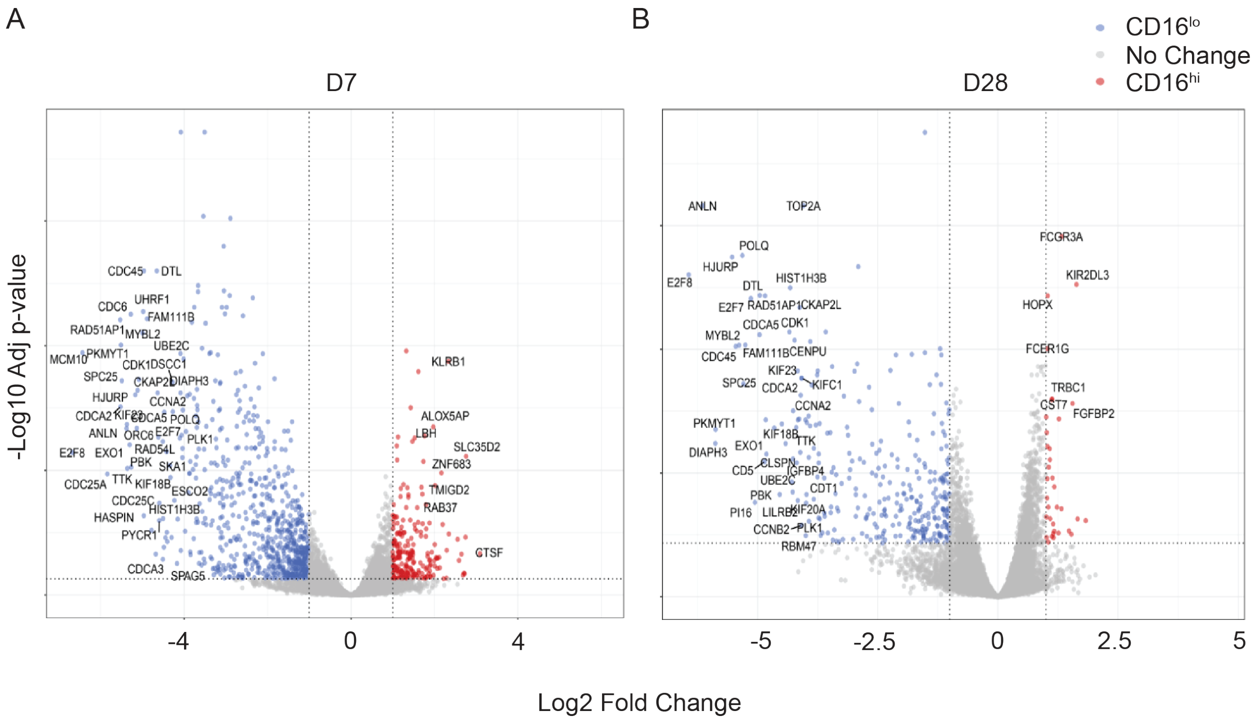


**Figure S7. Differential gene expression comparing CD16hi and CD16lo NK cells. A** Volcano plot showing genes more highly differentially expressed in CD16lo NK cells (blue) and genes more highly differentially expressed in CD16hi NK cells (red) on day +7 (D7). **B** Volcano plot showing genes more highly expressed in CD16lo NK cells (blue) and genes more highly expressed in CD16hi NK cells (red) on day +28 (D28).

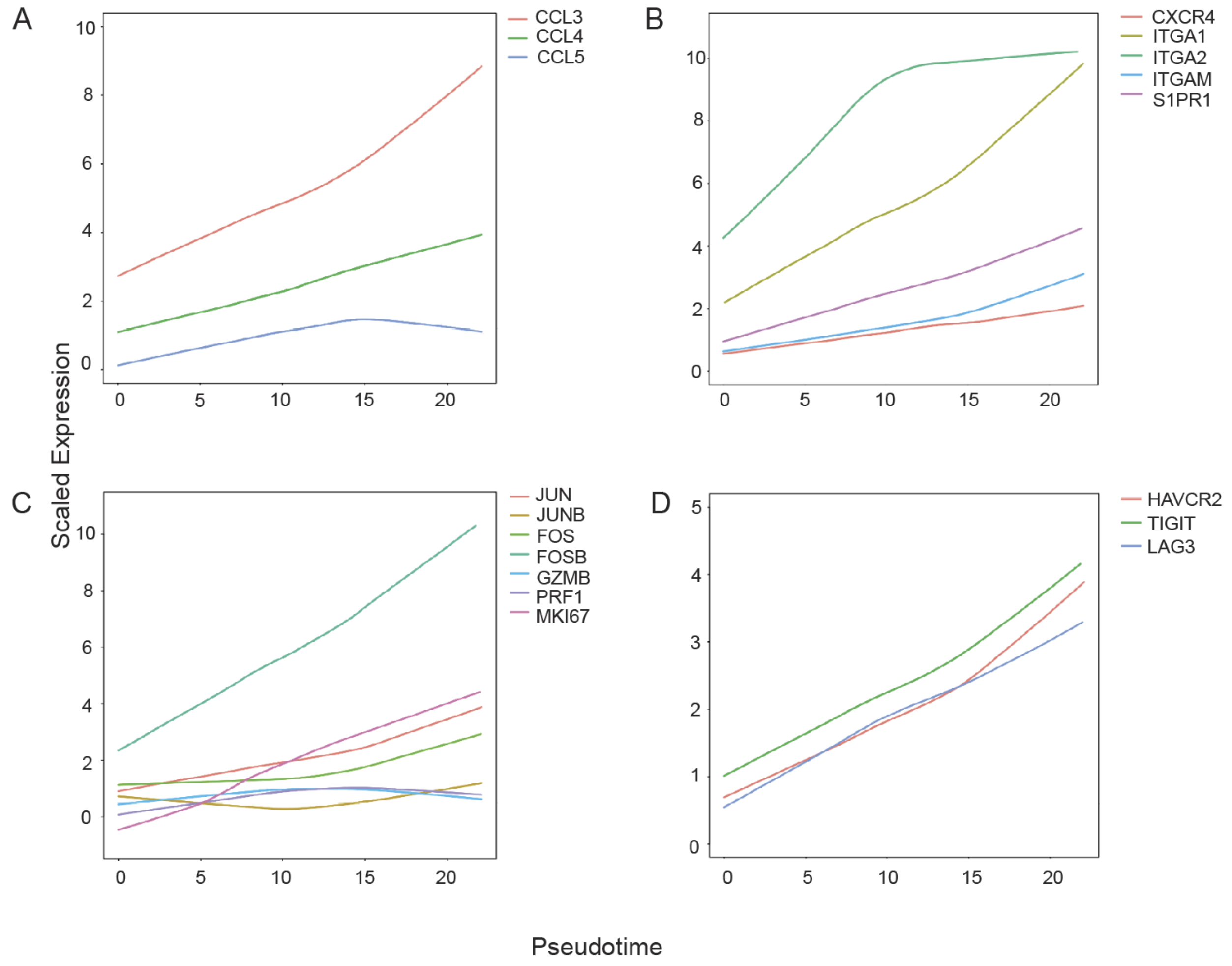


**Figure S8. Gene expression over pseudotime in NK cell clusters.** Pseudotime plots for chemokines (**A**), chemotaxis/adhesion (**B**), NK activation markers (**C**), and exhaustion markers (**D**).

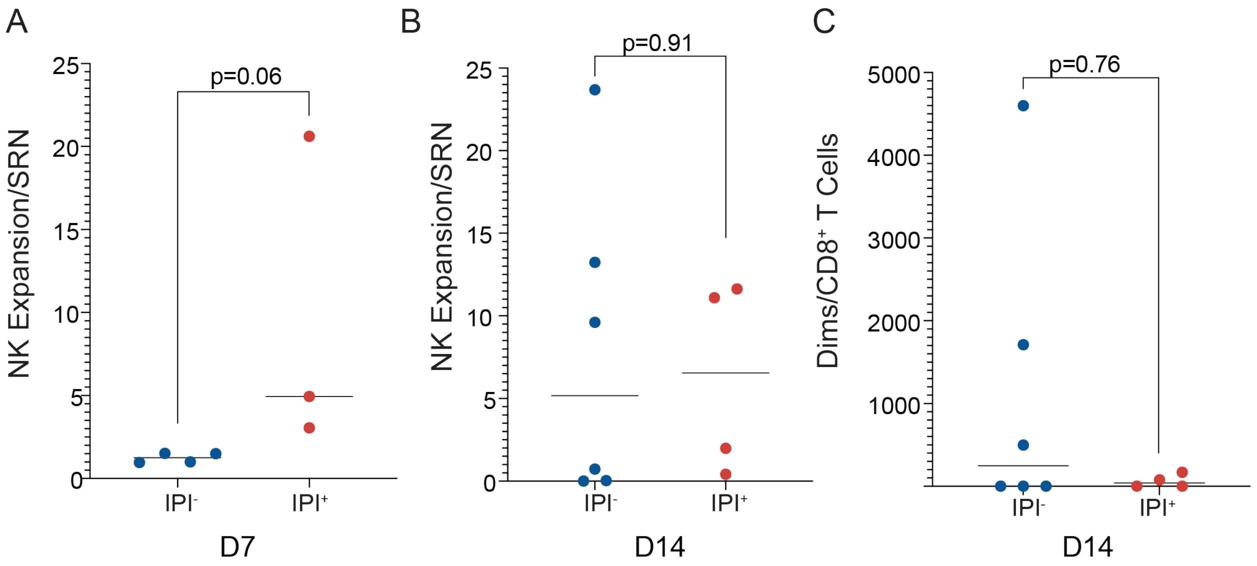


**Figure S9. NK cell expansion in the context of ipilimumab (IPI).** Fold expansion compared to the trial screening (SRN) quantity of NK cells (cells/uL) in the same patient, grouped by whether patients received IPI on day +7 (D7) in **A** and on day +14 (D14) in **B**. **C** Fold expansion of NK cells relative to SRN relative to fold expansion of CD8+ T cells compared to SRN in the same patient, grouped by whether patients received IPI on day +14.

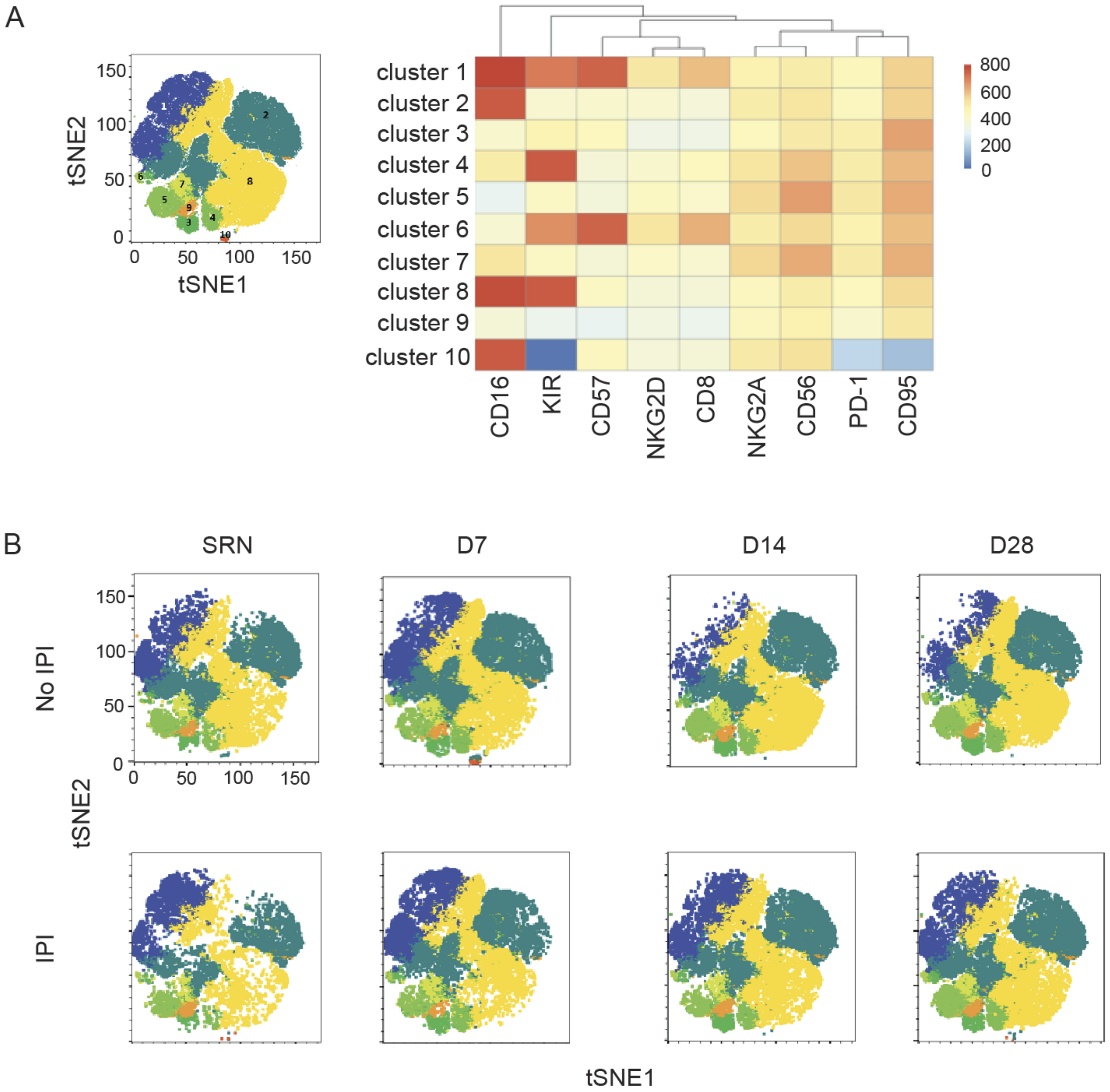


Figure S10. FlowSOM analysis of peripheral blood mononuclear cell populations following donor CIML NK infusion. A Clustering of NK cell populations on tSNE plot, with heatmap showing the numbered clusters based on measured markers. Highest expression of marker (red) is shown. B Longitudinal evaluation of NK cell clusters on tSNE plots from representative patients in each of the IPI treated and untreated groups.

**Figure S10. FlowSOM analysis of peripheral blood mononuclear cell populations following donor CIML NK infusion. A** Clustering of NK cell populations on tSNE plot, with heatmap showing the numbered clusters based on measured markers. Highest expression of marker (red) is shown. **B** Longitudinal evaluation of NK cell clusters on tSNE plots from representative patients in each of the IPI treated and untreated groups.

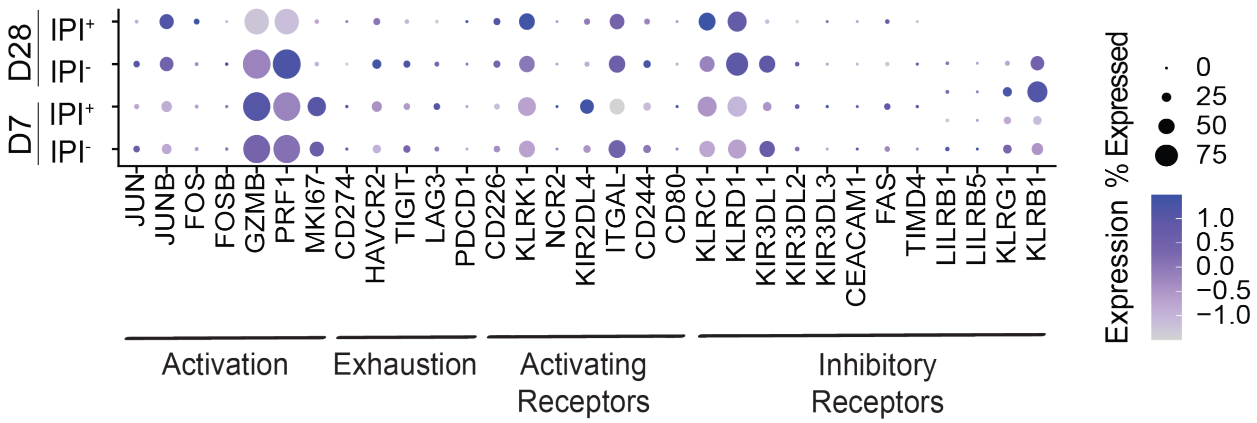


**Figure S11. Gene expression patterns in NK cells.** States of NK activation and exhaustion are shown on both day +7 (D7) and day +28 (D28), in patients who were not treated with IPI (n=2, IPI$^-$) and who were treated with IPI (n=2, IPI$^+$). NK activating and inhibitory receptors are also shown. The size of the dots reflects the number of cells expressing a gene, and the color reflects the degree of expression. P-values were calculated using Mann-Whitney U test and are detailed in Supplementary Table 14.

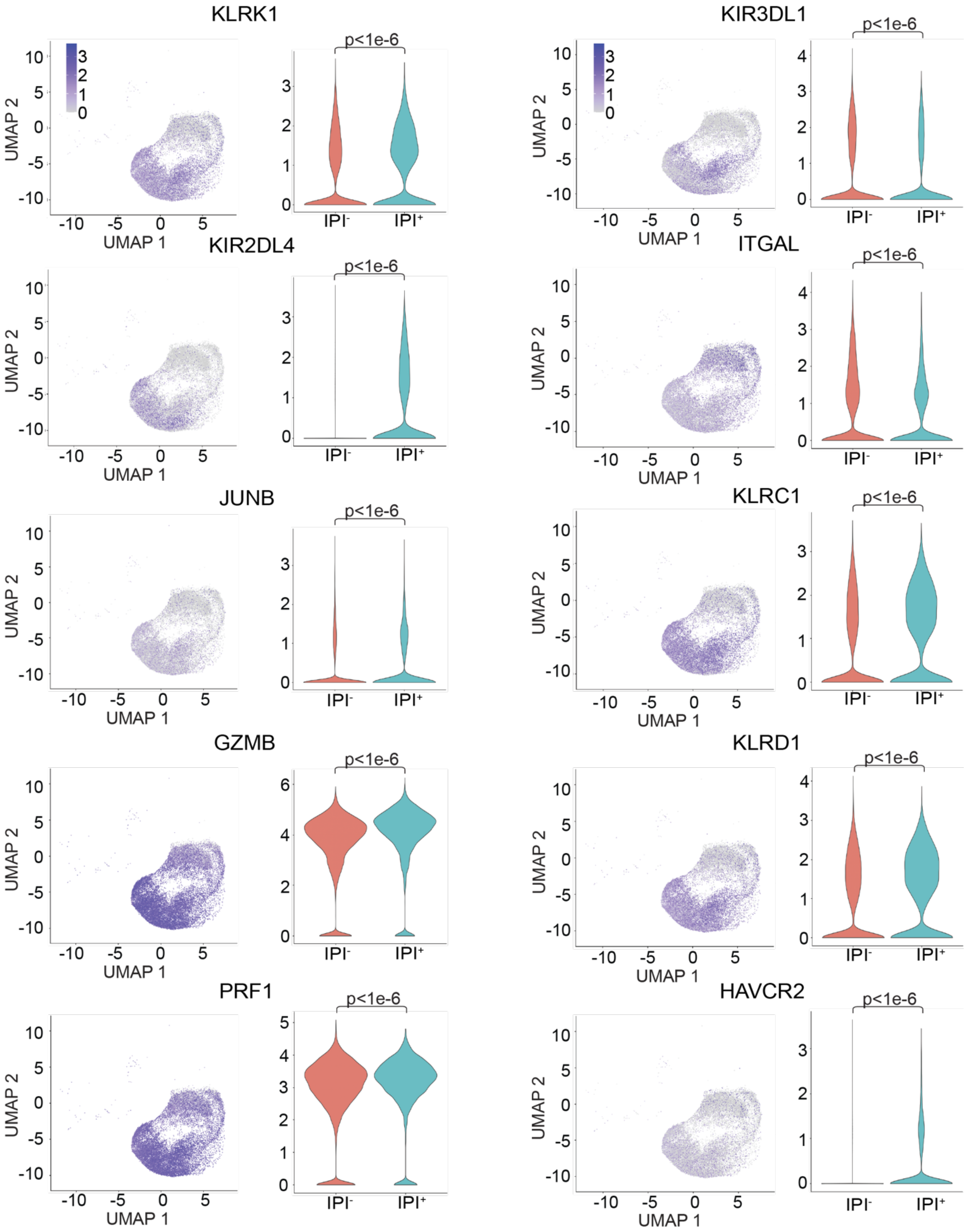


**Figure S12. Genes associated with NK cell activation and inhibition.** UMAP showing gene expression in NK cell clusters in all samples (n=4), with increased expression shown in purple. Concurrent gene expression shown in violin plots for each gene, comparing IPI$^{+}$ (n=2) and IPI$^{-}$ (n=2). Most differentially expressed genes are shown. Mann-Whitney U test, adjusted p-values.

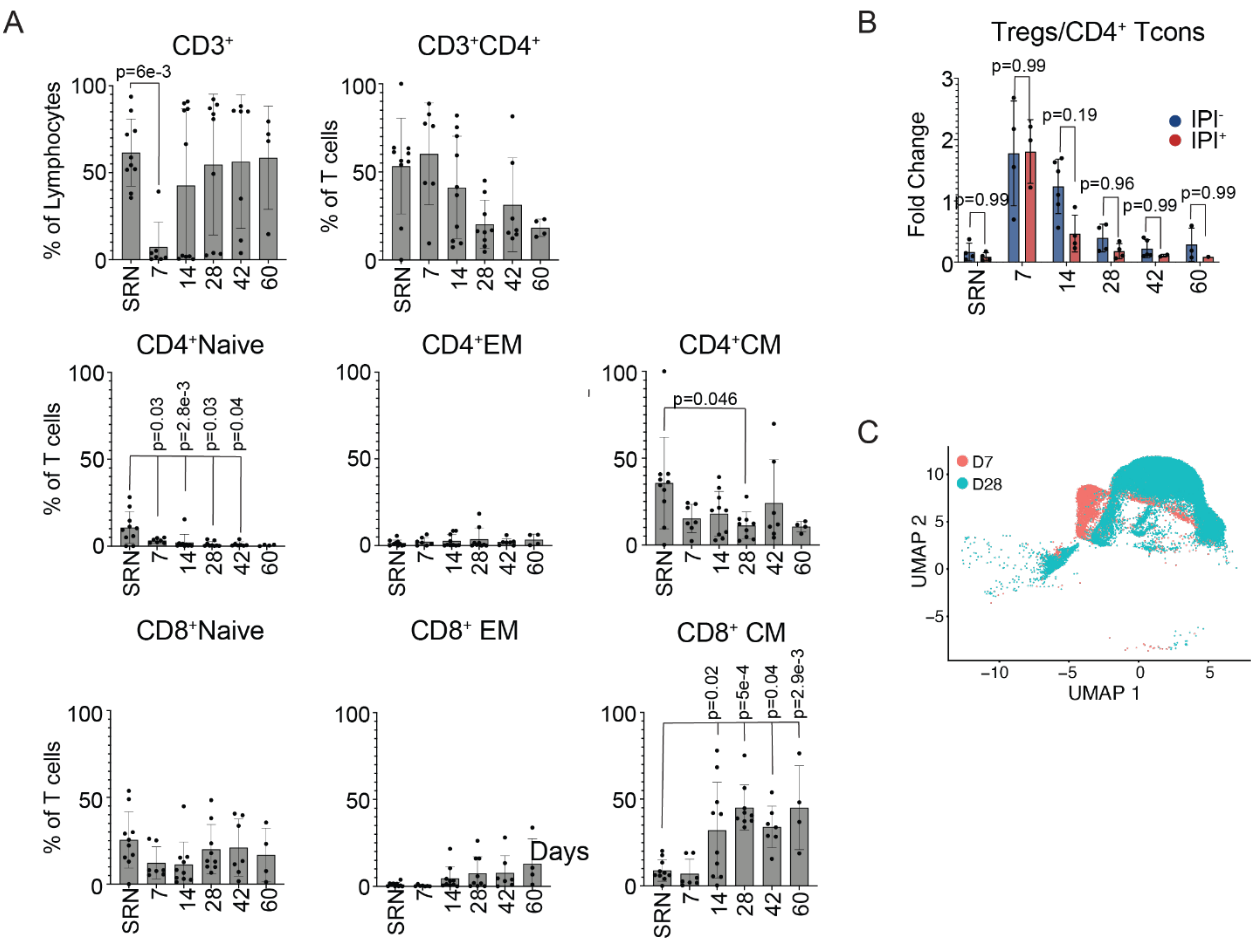


**Figure S13. T cell populations following NK cell infusion. A** Longitudinal evaluation of T cell populations evaluated with flow cytometry. Adjusted p-values for each time point comparison with SRN (trial screening time point) are determined using Dunnett's multiple comparisons test and detailed in Supplementary table 13, significant p-values are indicated. **B** Comparison of the Treg:Tcon in those treated without IPI (IPI$^{-}$, n=6) to those treated with IPI (IPI$^{+}$, n=4), Adjusted p-values per each time point were calculated with Šídák's multiple comparisons test. SRN: screening time point, D7: day +7, D14: day +14, D28: day +28 following CIML NK infusion. **C** T cell subpopulations as clustered with CITEseq on day +7 (D7) and day +28 (D28), n=4 at each time point.

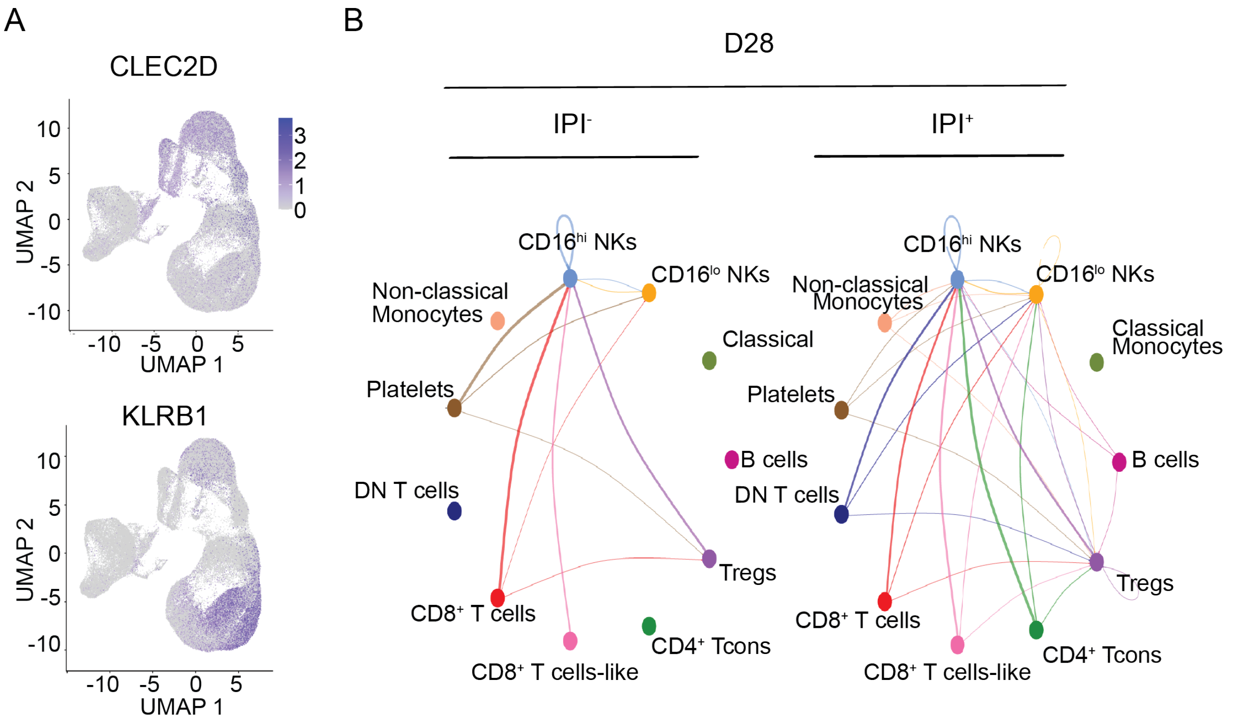


**Figure S14. Interaction analysis via the KLRB1-CLEC2D signaling axis.** Predicted cell-cell interaction with CellChat between T cell populations expressing the NK ligand CLEC2D and NK cells expressing its receptor KLRB1 on day +28 (D28) (**A**), with interaction maps showing both IPI$^-$ and IPI$^+$ contexts (**B**).

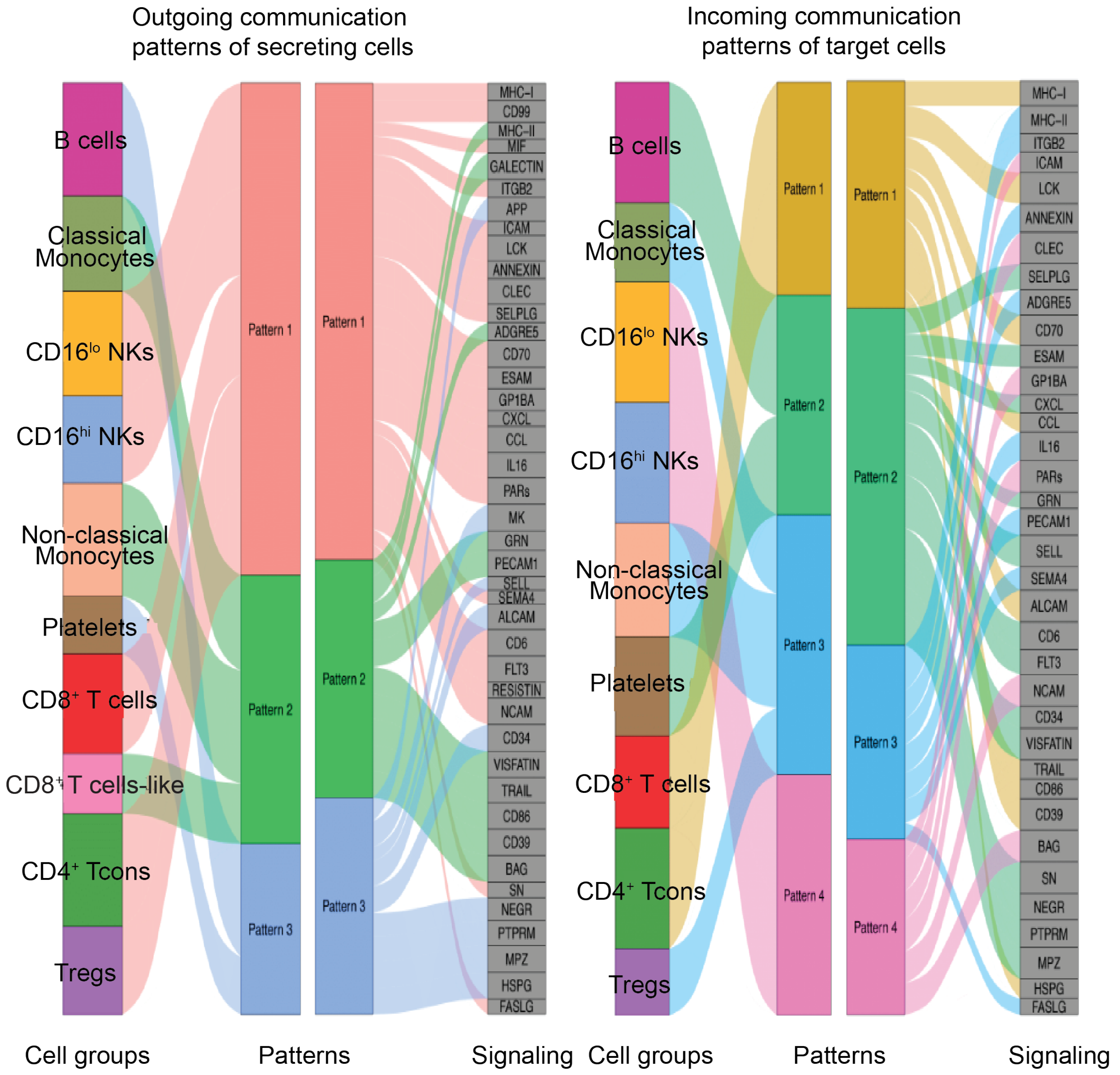


**Figure S15. River plot of cell-cell interaction analysis with CellChat.** River plots from CellChat analysis of CITEseq data from peripheral blood mononuclear cells on day +28 following donor NK cell infusion, showing both incoming and outgoing communication patterns in all cells.

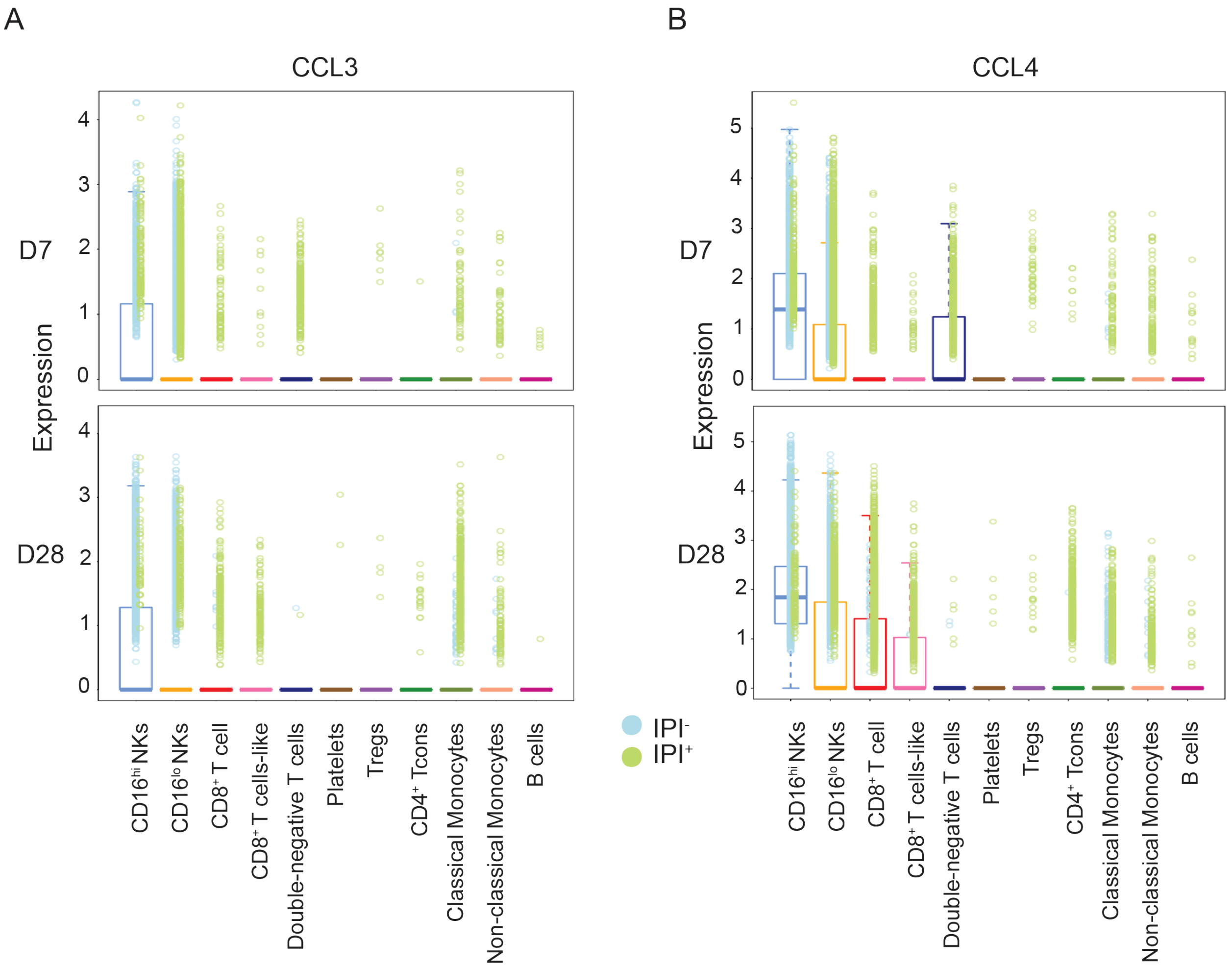


**Figure S16. Gene expression of chemotaxis molecules in peripheral blood mononuclear cell populations.** Scaled expression of chemotaxis genes CCL3 (**A**) and CCL4 (**B**) in each of the indicated cell subpopulations, showing expression of the indicated markers in patients who did not receive IPI ($IPI^-$, blue) and those who did ($IPI^+$, green) on day +7 (D7) and day +28 (D28). Expression is shown as boxplots, with the box showing median and quartiles. Where a box is not shown, the dots are outliers. P-values were calculated using Mann-Whitney U test and are detailed in **Supplementary Table 16**.

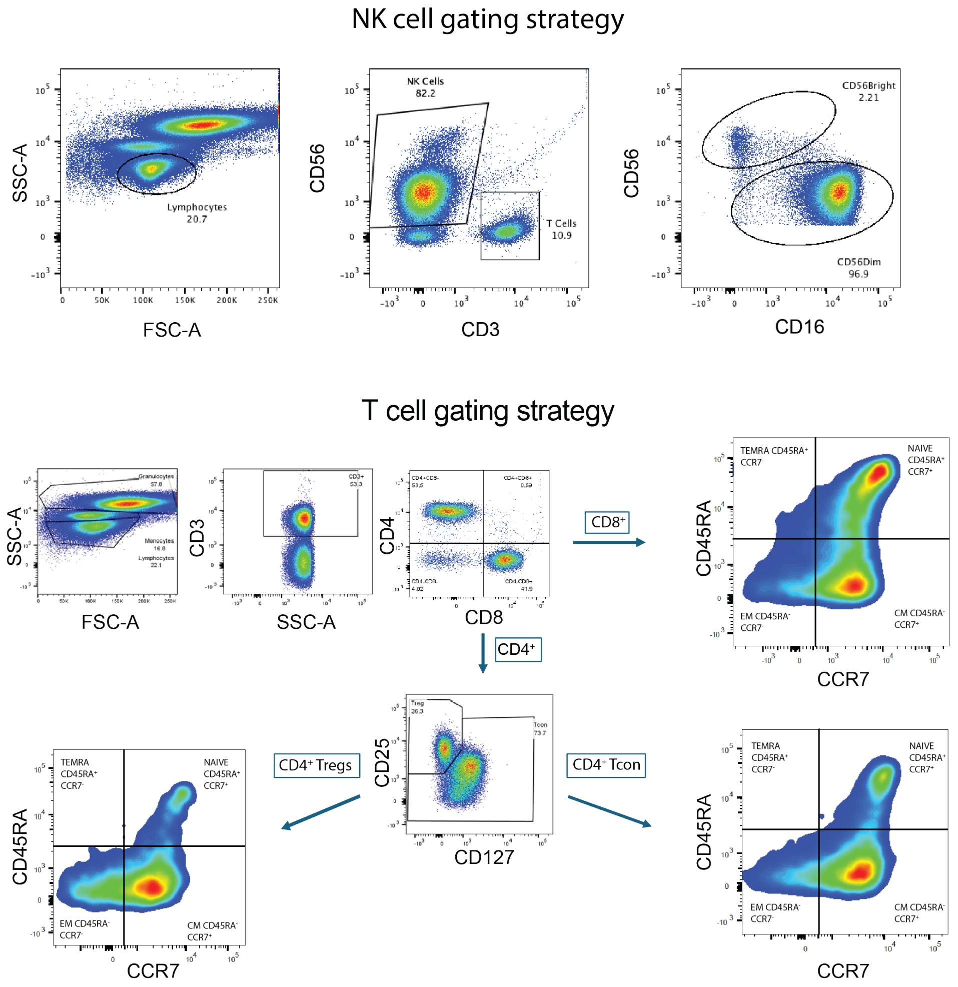


**Figure S17. Flow cytometry gating strategies for NK cells and T cells.** Representative gating strategies are shown from select samples and are consistent across all samples.

**Table S1. Clinical characteristics of patients enrolled on the phase I trial**

| Patient[¶] | Primary Diagnosis[A] | Initial Stage | Genetics of Disease[B] | Lines of prior therapy | CIML NK cell dose (cell/kg)[C] | Best Overall Response[D] | CRS[E] | Neurotox[F] | GVHD[G] |
|---|---|---|---|---|---|---|---|---|---|
| Patient 1 | OPC-HPV+ | T2N1-I | BCL2L1<br>MYC amplification<br>FGFR3 | 5 | $10x10^6$ | PR | Yes | No | No |
| Patient 2 | OPC-HPV+ | T2N1-I | STK11<br>FGFR deletion<br>PDCD1 mutation | 4 | $10x10^6$ | PD | Yes | No | No |
| Patient 3 | OPC-HPV+ | T2N1-I | CHEK2 loss<br>NF1<br>PTEN loss | 7 | $10x10^6$ | SD | Yes | No | No |
| Patient 4 | ACC-SBM | T2N0-II | NTRK3<br>RARA<br>ROS1 | 8 | $10x10^6$ | SD | No | No | No |
| Patient 5 | OPC-HPV+ | T2N0-I | CDK12<br>FANCM | 8 | $10x10^6$ | SD | Yes | No | No |
| Patient 6 | OPC/OC | T2N2a-IVA | MTUYH<br>RB1 rearranged<br>TERTp<br>TP53 | 5 | $7.45x10^6$ | SD | Yes | No | No |
| Patient 7 | OPC-HPV+ | T3N1-II | FBXW7 | 3 | $6.39x10^6$ | SD | Yes | No | No |

| | | | | | | | | | |
|---|---|---|---|---|---|---|---|---|---|
| Patient 8 | ACC-PNS | T4aN0-IVA | CHEK2<br>FGF10/14<br>NOTCH1<br>PDCD1LG2<br>PDGFRB<br>POLE<br>SMAD4<br>rearranged | 3 | $10x10^6$ | PD | No | No | No |
| Patient 9 | ACC-BOT | T4aN2b-IVA | MYB-NFIB<br>TP53<br>PIK3R1 | 7 | $7.67x10^6$ | SD | No | No | No |
| Patient 10 | OPC-HPV+ | T2N2-III | STK11<br>BRCA1 | 3 | $10x10^6$ | PD | No | No | No |
| Patient 11* | OC-RMT | T4aN2c-IVA | NOTCH1,<br>TERTp,<br>TP53 | 2 | NA | NA | NA | NA | NA |

[††] representativeness of study participants is detailed in Table S17
[A] OPC: oropharyngeal carcinoma, HPV: human papillomavirus, ACC: adenoid cystic carcinoma, SBM: submandibular gland, OC: oral cavity, PNS: paranasal sinus, BOT: base of tongue, RMT: retromolar trigone
[B] Determined with next-generation sequencing at the time of diagnosis
[C] Clinical trial treatment. Flu: fludarabine, Cy: Cyclophosphamide given prior to haploidentical donor CIML NK: cytokine-induced memory-like natural killer cell, IL-15sa: interleukin-15 superagonist, IPI: ipilimumab
[D] Investigator assessed best overall response using RECIST v1.1 on day +30 after CIML NK infusion. PR: partial response, PD: progressive disease, SD: stable disease.
[E] Cytokine release syndrome following clinical trial therapy, all were Grade 1
[F] Neurotoxicity, evaluated during the dose-limiting toxicity period following clinical trial therapy
[G] Graft-versus-host disease, evaluated during the dose-limiting toxicity period following clinical trial therapy
* This patient was enrolled on the study but died from disease progression during the lymphodepletion and did not receive donor CIML NK cells. NA: not available.

**Table S2**. Pleural fluid flow cytometry following CIML NK cell infusion in patient 3

| **Time after infusion** | **NK cell markers %*** | | **T cell makers %** | | **B cell markers %** | | **Malignant cells**** |
|---|---|---|---|---|---|---|---|
| Day +14‡ | | | | | | | None |
| | CD16 | 92 | CD3 | 1 | CD19 | 0 | |
| | CD56 | 98 | $CD3^{+}CD4^{+}$ | 1 | CD20 | 0 | |
| | CD57 | 22 | CD3+CD8+ | 0 | CD22 | 0 | |
| Day +28†† | | | | | | | None |
| | CD16 | 93 | CD3 | 3 | CD19 | 0 | |
| | CD56 | 95 | $CD3^{+}CD4^{+}$ | 0 | CD20 | 0 | |
| | CD57 | 51 | $CD3^{+}CD8^{+}$ | 2 | CD22 | 0 | |

*percent of total gated events on flow cytometry
** assessed with cytology
‡ Flow cytometric gating is by CD45 and side scatter on lymphoid cells with 92% of the total cells in the gate. Analysis of the right pleural fluid specimen demonstrates 1% of gated events (1% of total events) as $CD3^{+}$ T cells. NK cells expressing CD16 (majority positive), CD57 (subset), and CD56 comprise 98% of gated events (89% of total events).
†† Flow cytometric gating is by CD45 and side scatter on lymphoid cells with 33% of the total cells in the gate. Analysis of the right pleural fluid specimen demonstrates 3% of gated events (<1% of total events) as $CD3^{+}$ T cells. NK cells expressing CD16, CD57 (variable), and CD56 comprise 92% of gated events (21% of total events).

**Table S3. HLA typing of recipients and donors**

| Patient | Recipient HLA | Donor HLA | Donor Specific Antibody |
|---|---|---|---|
| Patient 1 | A*68:01, A*03:01<br>B*44:02, B*08:01<br>C*07:04, C*07:01<br>DRB1*15:01, DRB1*15:01<br>DQA1*01:02, DQA1*01:02<br>DQB1*06:02<br>DRB5*01:01, DRB5*01:01<br>DPB1*04:01, DPB1*04:01 | A*68:01, A*24:02<br>B*44:02, B*44:02<br>C*07:04, C*05:01<br>DRB1*15:01, DRB1*01:01<br>DQA1*01:02, DQA1*01:01<br>DQB1*06:02, DQB1*05:01<br>DRB5*01:01<br>DPB1*04:01, DPB1*03:01 | none |
| Patient 2 | A*02:01, A*11:01<br>B*44:02, B*55:01<br>C*05:01, C*03:03<br>DRB1*15:01, DRB1*04:07<br>DQA1*01:02, DQA1*03:03<br>DQB1*06:02, DQB1*03:01<br>DRB5*01:01, DRB4*01:03<br>DPB1*04:01, DPB1*03:01 | A*02:01, A*02:05<br>B*44:02, B*49:01<br>C*05:01, C*07:01<br>DRB1*15:01, DRB1*04:05<br>DQA1*01:02, DQA1*03:03<br>DQB1*06:02, DQB1*03:02<br>DRB5*01:01, DRB4*01:03<br>DPB1*04:01, DPB1*04:01 | none |
| Patient 3 | A*02:01<br>B*44:02<br>C*05:01<br>DRB1*15:01, DRB1*09:01<br>DQA1*01:02, DQA1*03:02<br>DQB1*06:02, DQB1*03:03<br>DRB5*01:01, DRB4*01:03<br>DPB1*04:01, DPB1*04:01 | A*02:01, A*68:01<br>B*44:02, B*46:01<br>C*05:01, C*01:02<br>DRB1*15:01, DRB1*09:01<br>DQA1*01:02, DQA1*03:02<br>DQB1*06:02, DQB1*03:03<br>DRB5*01:01, DRB4*01:03<br>DPB1*04:01, DPB1*13:01 | none |
| Patient 4 | A*33:01, A*68:01<br>B*14:02, B*51:01<br>C*08:02, C*14:02<br>DRB1*03:01, DRB1*09:01<br>DQA1*05:01, DQA1*03:02<br>DQB1*02:01, DQB1*03:03<br>DRB3*02:02, DRB4*01:03<br>DPB1*02:01, DPB1*14:01 | A*03:01, A*68:01<br>B*07:02, B*51:01<br>C*07:02, C*14:02<br>DRB1*15:01, DRB1*09:01<br>DQA1*01:02, DQA1*03:02<br>DQB1*06:02, DQB1*03:03<br>DRB5*01:01, DRB4*01:03<br>DPB1*04:01, DPB1*14:01 | B44, B45 |
| Patient 5 | A*02:01, A*03:01<br>B*40:01, B*15:01<br>C*03:04, C*07:02<br>DRB1*08:01, DRB1*04:01<br>DQA1*04:01, DQA1*03:01<br>DQB1*04:02, DQB1*03:02<br>DRB4*01:03<br>DPB1*03:01, DPB1*04:01 | A*01:01, A*03:01<br>B*08:01, B*15:01<br>C*07:01, C*07:02<br>DRB1*09:01, DRB1*04:01<br>DQA1*03:02, DQA1*03:01<br>DQB1*03:03, DQB1*03:02<br>DRB4*01:03<br>DPB1*04:02, DPB1*04:01 | A25, A32, B44 |

| | | | |
|---|---|---|---|
| Patient 6 | A*29:02, A*02:01<br>B*44:03, B*44:02<br>C*16:01, C*05:01<br>DRB1*07:01, DRB1*04:01<br>DQA1*02:01, DQA1*03:03<br>DQB1*02:02, DQB1*03:01<br>DRB4*01:01, DRB4*01:03<br>DPB1*11:01, DPB1*04:01 | A*29:02, A*03:01<br>B*44:03, B*47:01<br>C*16:01, C*06:02<br>DRB1*07:01<br>DQA1*02:01<br>DQB1*02:02<br>DRB4*01:01<br>DPB1*11:01, DPB1*06:01 | B67 |
| Patient 7 | A*01:01<br>B*15:220, B*08:01<br>C*12:03, C*07:01<br>DRB1*07:01, DRB1*03:01<br>DQA1*02:01, DQA1*05:01<br>DQB1*02:02, DQB1*02:01<br>DRB4*01:03, DRB3*01:01<br>DPB1*02:01, DPB1*01:01 | A*24:02, A*01:01<br>B*51:01, B*08:01<br>C*16:02, C*07:01<br>DRB1*13:01, DRB1*03:01<br>DQA1*01:03, DQA1*05:01<br>DQB1*06:03, DQB1*02:01&<br>DRB4*01:01&, DRB4*01:01&<br>DPB1*04:01, DPB1*01:01 | none |
| Patient 8 | A*02:01, A*31:01<br>B*15:01, B*35:02<br>C*03:04, C*04:01<br>DRB1*04:01, DRB1*04:03<br>DQA1*03:01<br>DQB1*03:02, DQB1*03:05<br>DRB4*01:03, DPB1*04:01 | A*02:01, A*26:01<br>B*15:01, B*38:01<br>C*03:04, C*12:03<br>DRB1*04:01, DRB1*12:03<br>DQA1*03:01<br>DQB1*03:02, DQB1*03:02<br>DRB4*01:03, DPB1*04:01 | A23, A24, A25, A32,<br>A66, B13, B27, B37,<br>**B38**, B44, B47, B49,<br>B51, B52, B53, B57,<br>B58, B59, B63, B76,<br>B77 |
| Patient 9 | A*33:03, A*68:02<br>B*44:03, B*53:01<br>C*04:01 DRB1*15:03&,<br>DRB1*13:03<br>DQA1*01:02&, DQA1*02:01<br>DQB1*06:02, DQB1*02:02&<br>DRB5*01:01, DRB3*02:02<br>DPB1*01:01, DPB1*17:01 | A*74:01, A*68:02<br>B*15:03, B*53:01<br>C*02:10, C*04:01<br>DRB1*11:01, DRB1*13:03<br>DQA1*05:05, DQA1*02:01<br>DQB1*03:19, DQB1*02:02<br>DRB3*02:02, DRB4*02:02<br>DPB1*01:01, DPB1*17:01 | B82 |
| Patient 10 | A*03:01, A*02:01<br>B*35:01, B*40:01<br>C*04:01, C*03:04<br>DRB1*01:01, DRB1*08:01&<br>DQA1*01:01, DQA1*04:02<br>DQB1*05:01&, DQB1*04:02<br>DPB1*01:01, DPB1*03:01 | A*03:01, A*02:01<br>B*35:01, B*44:02<br>C*04:01, C*05:01<br>DRB1*01:01, DRB1*04:01<br>DQA1*01:01, DQA1*03:02<br>DQB1*05:01&, DQB1*03:01&<br>DRB4*01:03<br>DPB1*01:01, DPB1*04:01 | none |

**Table S4. Distribution of FlowSOM defined NK cell metaclusters in responders and non-responders**

| FlowSOM Metaclusters* | SRN | | D7 | | D14 | | D28 | |
|---|---|---|---|---|---|---|---|---|
| | Non Resp (n=5) (%) | Resp (n=4) (%) | Non Resp (n=5) (%) | Resp (n=4) (%) | Non Resp (n=5) (%) | Resp (n=4) (%) | Non Resp (n=5) (%) | Resp (n=4) (%) |
| **Cluster 1** | 24.1 | 24.5 | 31.7 | 20 | **1.23** | **11.74** | **0.64** | **18.0** |
| **Cluster 2** | 29.6 | 32.7 | 29.7 | 41.6 | 30.3 | 20.4 | 37.9 | 46.3 |
| **Cluster 3** | 1.49 | 1.2 | 3.75 | 1.66 | **15.9** | **1.48** | 6.18 | 8.46 |
| **Cluster 4** | 1.40 | 0.67 | 5 | 3.68 | 2.82 | 2.84 | 2.23 | 0.031 |
| **Cluster 5** | 4.75 | 2.12 | 4.32 | 1.4 | **26.8** | **0.66** | **14.5** | **1.30** |
| **Cluster 6** | 0.075 | 0.045 | 2.21 | 0.51 | 0.21 | 0.033 | 0.33 | 1.30 |
| **Cluster 7** | 1.07 | 1.04 | 3.68 | 1.92 | **13.4** | **0.73** | **6.94** | **0.61** |
| **Cluster 8** | 34.6 | 36.7 | 19.1 | 26.3 | **5.78** | **61.6** | 29.7 | 18.4 |
| **Cluster 9** | 2.89 | 0.94 | 0.59 | 0.45 | 3.52 | 0.49 | 1.61 | 5.55 |
| **Cluster 10** | 0.0 | 0.025 | 0 | 2.44 | 0 | 0 | 0 | 0.01 |

*Clusters correspond to those defined in Figure S10
SRN: trial screening time point, D7: day +7, D14: day +14, D28: day +28, Non Resp: non-responder, Resp: responder based on RECIST v1.1 criteria.

**Table S5. Antibody panels for flow cytometry**

| Antibody | Fluorochrome | Clone | Amount (uL) | Vendor | Catalog # |
|---|---|---|---|---|---|
| **Flow Analysis** | | | | | |
| CD25 | FITC | m-A251 | 1.0 | BioLegend | 356105 |
| PD-1 | PE | J105 | 2.0 | Invitrogen | 12-2799-42 |
| CD95 | PeCF594 | DX2 | 1.0 | BioLegend | 305633 |
| TCR a/b | PeCy 7 | IP26 | 1.0 | BioLegend | 306719 |
| CD127 | APC | eBioRDR5 | 5.0 | Invitrogen | 17-1278-42 |
| CD8 | Alexa 700 | RPA-T8 | 1.0 | BioLegend | 301027 |
| TCR g/d | APC-Vio 770 | 11F2 | 2.0 | Miltenyi | 130-113-501 |
| TIM 3 | BV421 | F38-2E2 | 5.0 | BioLegend | 345007 |
| CD4 | BV510 | RPA-T4 | 2.0 | BioLegend | 300545 |
| CD45RA | BV605 | HL100 | 2.0 | BioLegend | 304133 |
| CD6 | BV650 | M-T605 | 2.0 | BD | 743448 |
| CCR7 | BV711 | G043-H7 | 5.0 | BioLegend | 353227 |
| CD3 | BV786 | UCHT1 | 2.0 | BioLegend | 300472 |
| CD16 | FITC | 3G8 | 2.0 | BD | 555406 |
| NKG2D | APC | 1D11 | 10.0 | BD | 558071 |
| CD3 | BV786 | UCHT1 | 2.0 | BioLegend | 300472 |
| CD56 | BV605 | NCAM | 1.0 | BioLegend | 318333 |
| CD57 | PerCP Cy5.5 | HNK-1 | 2.0 | BioLegend | 359621 |
| NKG2A | BV421 | 131411 | 1.0 | BD | 747924 |
| CD158a | PeCy7 | DX27 | 1.0 | BioLegend | 312609 |
| CD158b | PeCy7 | HP-MA4 | 1.0 | BioLegend | 339511 |
| CD158e | PeCy7 | DX9 | 1.0 | BioLegend | 312720 |
| **Functional Assays** | | | | | |
| CD56 | PE | HCD56 | 1.0 | BioLegend | 318306 |
| CD3 | BV510 | HIT3a | 1.0 | BD | 564713 |
| CD16 | APC-Cy7 | 3G8 | 1.0 | BioLegend | 302018 |
| CD107a | APC | H4A3 | 1.0 | BioLegend | 328620 |
| IFNg | PE-Cy7 | B27 | 1.0 | BD | 557643 |

Beckton Dickinson and Company, NJ, USA; Biolegend, CA, USA; Invitrogen, MA, USA; Miltenyi Biotec, North Rhine-Westphalia , Germany

**Table S6. Panel of CITEseq antibodies (TotalSeq™-C Human Universal Cocktail, V1.0, Biolegend supplemented with antibodies against NKG2A,NKG2C,NKp30) .**

| DNA_ID | Description | Clone | Barcode |
|---|---|---|---|
| C0006 | anti-human CD86 | IT2.2 | GTCTTTGTCAGTGCA |
| C0007 | anti-human CD274 (B7-H1, PD-L1) | 29E.2A3 | GTTGTCCGACAATAC |
| C0020 | anti-human CD270 (HVEM, TR2) | 122 | TGATAGAAACAGACC |
| C0023 | anti-human CD155 (PVR) | SKII.4 | ATCACATCGTTGCCA |
| C0024 | anti-human CD112 (Nectin-2) | TX31 | AACCTTCCGTCTAAG |
| C0026 | anti-human CD47 | CC2C6 | GCATTCTGTCACCTA |
| C0029 | anti-human CD48 | BJ40 | CTACGACGTAGAAGA |
| C0031 | anti-human CD40 | 5C3 | CTCAGATGGAGTATG |
| C0032 | anti-human CD154 | 24-31 | GCTAGATAGATGCAA |
| C0033 | anti-human CD52 | HI186 | CTTTGTACGAGCAAA |
| C0034 | anti-human CD3 | UCHT1 | CTCATTGTAACTCCT |
| C0046 | anti-human CD8 | SK1 | GCGCAACTTGATGAT |
| C0047 | anti-human CD56 | 5.1H11 | TCCTTTCCTGATAGG |
| C0050 | anti-human CD19 | HIB19 | CTGGGCAATTACTCG |
| C0052 | anti-human CD33 | P67.6 | TAACTCAGGGCCTAT |
| C0053 | anti-human CD11c | S-HCL-3 | TACGCCTATAACTTG |
| C0058 | anti-human HLA-A,B,C | W6/32 | TATGCGAGGCTTATC |
| C0063 | anti-human CD45RA | HI100 | TCAATCCTTCCGCTT |
| C0064 | anti-human CD123 | 6H6 | CTTCACTCTGTCAGG |
| C0066 | anti-human CD7 | CD7-6B7 | TGGATTCCCGGACTT |
| C0068 | anti-human CD105 | 43A3 | ATCGTCGAGAGCTAG |
| C0070 | anti-human/mouse CD49f | GoH3 | TTCCGAGGATGATCT |
| C0071 | anti-human CD194 (CCR4) | L291H4 | AGCTTACCTGCACGA |
| C0072 | anti-human CD4 | RPA-T4 | TGTTCCCGCTCAACT |
| C0073 | anti-mouse/human CD44 | IM7 | TGGCTTCAGGTCCTA |
| C0081 | anti-human CD14 | M5E2 | TCTCAGACCTCCGTA |
| C0083 | anti-human CD16 | 3G8 | AAGTTCACTCTTTGC |
| C0085 | anti-human CD25 | BC96 | TTTGTCCTGTACGCC |
| C0087 | anti-human CD45RO | UCHL1 | CTCCGAATCATGTTG |
| C0088 | anti-human CD279 | EH12.2H7 | ACAGCGCCGTATTTA |
| C0089 | anti-human TIGIT (VSTM3) | A15153G | TTGCTTACCGCCAGA |
| C0090 | Mouse IgG1, κ isotype Ctrl | MOPC-21 | GCCGGACGACATTAA |
| C0091 | Mouse IgG2a, κ isotype Ctrl | MOPC-173 | CTCCTACCTAAACTG |
| C0092 | Mouse IgG2b, κ isotype Ctrl | MPC-11 | ATATGTATCACGCGA |
| C0095 | Rat IgG2b, κ Isotype Ctrl | RTK4530 | GATTCTTGACGACCT |
| C0100 | anti-human CD20 | 2H7 | TTCTGGGTCCCTAGA |
| C0101 | anti-human CD335 (NKp46) | 9E2 | ACAATTTGAACAGCG |
| C0124 | anti-human CD31 | WM59 | ACCTTTATGCCACGG |

| | | | |
|---|---|---|---|
| C0134 | anti-human CD146 | P1H12 | CCTTGGATAACATCA |
| C0136 | anti-human IgM | MHM-88 | TAGCGAGCCCGTATA |
| C0138 | anti-human CD5 | UCHT2 | CATTAACGGGATGCC |
| C0140 | anti-human CD183 (CXCR3) | G025H7 | GCGATGGTAGATTAT |
| C0141 | anti-human CD195 (CCR5) | J418F1 | CCAAAGTAAGAGCCA |
| C0142 | anti-human CD32 | FUN-2 | GCTTCCGAATTACCG |
| C0143 | anti-human CD196 (CCR6) | G034E3 | GATCCCTTTGTCACT |
| C0144 | anti-human CD185 (CXCR5) | J252D4 | AATTCAACCGTCGCC |
| C0145 | anti-human CD103 (Integrin αE) | Ber-ACT8 | GACCTCATTGTGAAT |
| C0146 | anti-human CD69 | FN50 | GTCTCTTGGCTTAAA |
| C0147 | anti-human CD62L | DREG-56 | GTCCCTGCAACTTGA |
| C0149 | anti-human CD161 | HP-3G10 | GTACGCAGTCCTTCT |
| C0151 | anti-human CD152 (CTLA-4) | BNI3 | ATGGTTCACGTAATC |
| C0152 | anti-human CD223 (LAG-3) | 11C3C65 | CATTTGTCTGCCGGT |
| C0153 | anti-human KLRG1 (MAFA) | SA231A2 | CTTATTTCCTGCCCT |
| C0154 | anti-human CD27 | O323 | GCACTCCTGCATGTA |
| C0155 | anti-human CD107a (LAMP-1) | H4A3 | CAGCCCACTGCAATA |
| C0156 | anti-human CD95 (Fas) | DX2 | CCAGCTCATTAGAGC |
| C0158 | anti-human CD134 (OX40) | Ber-ACT35 (ACT35) | AACCCACCGTTGTTA |
| C0159 | anti-human HLA-DR | L243 | AATAGCGAGCAAGTA |
| C0160 | anti-human CD1c | L161 | GAGCTACTTCACTCG |
| C0161 | anti-human CD11b | ICRF44 | GACAAGTGATCTGCA |
| C0162 | anti-human CD64 | 10.1 | AAGTATGCCCTACGA |
| C0163 | anti-human CD141 (Thrombomodulin) | M80 | GGATAACCGCGCTTT |
| C0164 | anti-human CD1d | 51.1 | TCGAGTCGCTTATCA |
| C0165 | anti-human CD314 (NKG2D) | 1D11 | CGTGTTTGTTCCTCA |
| C0167 | anti-human CD35 | E11 | ACTTCCGTCGATCTT |
| C0168 | anti-human CD57 Recombinant | QA17A04 | AACTCCCTATGGAGG |
| C0170 | anti-human CD272 (BTLA) | MIH26 | GTTATTGGACTAAGG |
| C0171 | anti-human/mouse/rat CD278 (ICOS) | C398.4A | CGCGCACCCATTAAA |
| C0174 | anti-human CD58 (LFA-3) | TS2/9 | GTTCCTATGGACGAC |
| C0176 | anti-human CD39 | A1 | TTACCTGGTATCCGT |
| C0179 | anti-human CX3CR1 | K0124E1 | AGTATCGTCTCTGGG |
| C0180 | anti-human CD24 | ML5 | AGATTCCTTCGTGTT |
| C0181 | anti-human CD21 | Bu32 | AACCTAGTAGTTCGG |
| C0185 | anti-human CD11a | TS2/4 | TATATCCTTGTGAGC |
| C0187 | anti-human CD79b (Igβ) | CB3-1 | ATTCTTCAACCGAAG |
| C0189 | anti-human CD244 (2B4) | C1.7 | TCGCTTGGATGGTAG |
| C0206 | anti-human CD169 (Sialoadhesin, Siglec-1) | 7-239 | TACTCAGCGTGTTTG |
| C0214 | anti-human/mouse integrin β7 | FIB504 | TCCTTGGATGTACCG |
| C0215 | anti-human CD268 (BAFF-R) | 11C1 | CGAAGTCGATCCGTA |

| | | | |
|---|---|---|---|
| C0216 | anti-human CD42b | HIP1 | TCCTAGTACCGAAGT |
| C0217 | anti-human CD54 | HA58 | CTGATAGACTTGAGT |
| C0218 | anti-human CD62P (P-Selectin) | AK4 | CCTTCCGTATCCCTT |
| C0219 | anti-human CD119 (IFN-γ R α chain) | GIR-208 | TGTGTATTCCCTTGT |
| C0224 | anti-human TCR α/β | IP26 | CGTAACGTAGAGCGA |
| C0236 | Rat IgG1, κ isotype Ctrl | RTK2071 | ATCAGATGCCCTCAT |
| C0238 | Rat IgG2a, κ Isotype Ctrl | RTK2758 | AAGTCAGGTTCGTTT |
| C0241 | Armenian Hamster IgG Isotype Ctrl | HTK888 | CCTGTCATTAAGACT |
| C0246 | anti-human CD122 (IL-2Rβ) | TU27 | TCATTTCCTCCGATT |
| C0247 | anti-human CD267 (TACI) | 1A1 | AGTGATGGAGCGAAC |
| C0352 | anti-human FcεRIα | AER-37 (CRA-1) | CTCGTTTCCGTATCG |
| C0353 | anti-human CD41 | HIP8 | ACGTTGTGGCCTTGT |
| C0355 | anti-human CD137 (4-1BB) | 4B4-1 | CAGTAAGTTCGGGAC |
| C0358 | anti-human CD163 | GHI/61 | GCTTCTCCTTCCTTA |
| C0359 | anti-human CD83 | HB15e | CCACTCATTTCCGGT |
| C0363 | anti-human CD124 (IL-4Rα) | G077F6 | CCGTCCTGATAGATG |
| C0364 | anti-human CD13 | WM15 | TTTCAACGCCCTTTC |
| C0367 | anti-human CD2 | TS1/8 | TACGATTTGTCAGGG |
| C0368 | anti-human CD226 (DNAM-1) | 11A8 | TCTCAGTGTTTGTGG |
| C0369 | anti-human CD29 | TS2/16 | GTATTCCCTCAGTCA |
| C0370 | anti-human CD303 (BDCA-2) | 201A | GAGATGTCCGAATTT |
| C0371 | anti-human CD49b | P1E6-C5 | GCTTTCTTCAGTATG |
| C0373 | anti-human CD81 (TAPA-1) | 5A6 | GTATCCTTCCTTGGC |
| C0384 | anti-human IgD | IA6-2 | CAGTCTCCGTAGAGT |
| C0385 | anti-human CD18 | TS1/18 | TATTGGGACACTTCT |
| C0386 | anti-human CD28 | CD28.2 | TGAGAACGACCCTAA |
| C0389 | anti-human CD38 | HIT2 | TGTACCCGCTTGTGA |
| C0390 | anti-human CD127 (IL-7Rα) | A019D5 | GTGTGTTGTCCTATG |
| C0391 | anti-human CD45 | HI30 | TGCAATTACCCGGAT |
| C0393 | anti-human CD22 | S-HCL-1 | GGGTTGTTGTCTTTG |
| C0394 | anti-human CD71 | CY1G4 | CCGTGTTCCTCATTA |
| C0396 | anti-human CD26 | BA5b | GGTGGCTAGATAATG |
| C0407 | anti-human CD36 | 5-271 | TTCTTTGCCTTGCCA |
| C0420 | anti-human CD158 (KIR2DL1/S1/S3/S5) | HP-MA4 | TATCAACCAACGCTT |
| C0575 | anti-human CD49a | TS2/7 | ACTGATGGACTCAGA |
| C0576 | anti-human CD49d | 9F10 | CCATTCAACTTCCGG |
| C0577 | anti-human CD73 (Ecto-5'-nucleotidase) | AD2 | CAGTTCCTCAGTTCG |
| C0581 | anti-human TCR Vα7.2 | 3C10 | TACGAGCAGTATTCA |
| C0582 | anti-human TCR Vδ2 | B6 | TCAGTCAGATGGTAT |
| C0591 | anti-human LOX-1 | 15C4 | ACCCTTTACCGAATA |

| | | | |
|---|---|---|---|
| C0592 | anti-human CD158b (KIR2DL2/L3, NKAT2) | DX27 | GACCCGTAGTTTGAT |
| C0599 | anti-human CD158e1 (KIR3DL1, NKB1) | DX9 | GGACGCTTTCCTTGA |
| C0830 | anti-human CD319 (CRACC) | 162.1 | AGTATGCCATGTCTT |
| C0845 | anti-human CD99 | 3B2/TA8 | ACCCGTCCCTAAGAA |
| C0853 | anti-human CLEC12A | 50C1 | CATTAGAGTCTGCCA |
| C0864 | anti-human CD352 (NTB-A) | NT-7 | AGTTTCCACTCAGGC |
| C0867 | anti-human CD94 | DX22 | CTTTCCGGTCCTACA |
| C0894 | anti-human Ig light chain κ | MHK-49 | AGCTCAGCCAGTATG |
| C0896 | anti-human CD85j (ILT2) | GHI/75 | CCTTGTGAGGCTATG |
| C0897 | anti-human CD23 | EBVCS-5 | TCTGTATAACCGTCT |
| C0898 | anti-human Ig light chain λ | MHL-38 | CAGCCAGTAAGTCAC |
| C0902 | anti-human CD328 (Siglec-7) | 6-434 | CTTAGCATTTCACTG |
| C0912 | anti-human GPR56 | CG4 | GCCTAGTTTCCGTTT |
| C0918 | anti-human HLA-E | 3D12 | GAGTCGAGAAATCAT |
| C0920 | anti-human CD82 | ASL-24 | TCCCACTTCCGCTTT |
| C0944 | anti-human CD101 (BB27) | BB27 | CTACTTCCCTGTCAA |
| C1046 | anti-human CD88 (C5aR) | S5/1 | GCCGCATGAGAAACA |
| C1052 | anti-human CD224 | KF29 | CTGATGAGATGTCAG |
| C1374 | anti-human NKG2A | S19004C | CAACTCCTGGGACTT |
| C1375 | anti-human NKG2C | S19005E | ATTCCTGCGCCTAAA |
| C0801 | anti-human NKp30 | P30-15 | AAAGTCACTCTGCCG |

**Table S7. Gene lists used in the analysis**

| Categories | Gene list |
| --- | --- |
| NK cell activation | JUN, JUNB, FOS, FOSB, GZMB, PRF1, MKI67 |
| NK cell activating receptors | CD226, KIR3DS1, KLRK1, NCR2, KIRDS4, ITGAL, KIR2DL4 |
| NK cell inhibitory receptors | KLRC1, KIR3DL1, KIR3DL2, KIR3DL3, LILRB1, LILRB5, KLRG1, KLRB1 |
| NK exhaustion | HAVCR2, TIGIT, LAG3, PDCD1 |
| NK cell ligands | HLA-E, LGALS9, PVR, NECTIN2, CD274, PDCD1LG2, CEACAM1, CEACAM5, MICA, MICB, ULBP1, ULBP2, ULBP3, RAET1E, KMT2E, NCR3LG1, HLA-G, HLA-A, HLA-C, ICAM1, CD48, S100A8, S100A9, CDH2, CDH1, CDH4, CLEC2D |

**Table S8. The panel of signaling molecules for measurement in patient plasma.**

| Analyte | Uniprot ID |
|---|---|
| IL8 | P10145 |
| TNFRSF9 | Q07011 |
| TIE2 | Q02763 |
| MCP-3 | P80098 |
| CD40-L | P29965 |
| IL-1 alpha | P01583 |
| CD244 | Q9BZW8 |
| EGF | P01133 |
| ANGPT1 | Q15389 |
| IL7 | P13232 |
| PGF | P49763 |
| IL6 | P05231 |
| ADGRG1 | Q9Y653 |
| MCP-1 | P13500 |
| CRTAM | O95727 |
| CXCL11 | O14625 |
| MCP-4 | Q99616 |
| TRAIL | P50591 |
| FGF2 | P09038 |
| CXCL9 | Q07325 |
| CD8A | P01732 |
| CAIX | Q16790 |
| MUC-16 | Q8WXI7 |
| ADA | P00813 |
| CD4 | P01730 |
| NOS3 | P29474 |
| IL2 | P60568 |
| Gal-9 | O00182 |
| VEGFR-2 | P35968 |
| CD40 | P25942 |
| IL18 | Q14116 |
| GZMH | P20718 |
| KIR3DL1 | P43629 |
| LAP TGF-beta-1 | P01137 |
| CXCL1 | P09341 |
| TNFSF14 | O43557 |
| IL33 | O95760 |
| TWEAK | O43508 |

| | |
|---|---|
| PDGF subunit B | P01127 |
| PDCD1 | Q15116 |
| FASLG | P48023 |
| CD28 | P10747 |
| CCL19 | Q99731 |
| MCP-2 | P80075 |
| CCL4 | P13236 |
| IL15 | P40933 |
| Gal-1 | P09382 |
| PD-L1 | Q9NZQ7 |
| CD27 | P26842 |
| CXCL5 | P42830 |
| IL5 | P05113 |
| HGF | P14210 |
| GZMA | P12544 |
| HO-1 | P09601 |
| CX3CL1 | P78423 |
| CXCL10 | P02778 |
| CD70 | P32970 |
| IL10 | P22301 |
| TNFRSF12A | Q9NP84 |
| CCL23 | P55773 |
| CD5 | P06127 |
| CCL3 | P10147 |
| MMP7 | P09237 |
| ARG1 | P05089 |
| NCR1 | O76036 |
| DCN | P07585 |
| TNFRSF21 | O75509 |
| TNFRSF4 | P43489 |
| MIC-A/B | Q29983,Q29980 |
| CCL17 | Q92583 |
| ANGPT2 | O15123 |
| PTN | P21246 |
| CXCL12 | P48061 |
| IFN-gamma | P01579 |
| LAMP3 | Q9UQV4 |
| CASP-8 | Q14790 |
| ICOSLG | O75144 |
| MMP12 | P39900 |
| CXCL13 | O43927 |

| | |
|---|---|
| PD-L2 | Q9BQ51 |
| VEGFA | P15692 |
| IL4 | P05112 |
| LAG3 | P18627 |
| IL12RB1 | P42701 |
| IL13 | P35225 |
| CCL20 | P78556 |
| TNF | P01375 |
| KLRD1 | Q13241 |
| GZMB | P10144 |
| CD83 | Q01151 |
| IL12 | P29459,P29460 |
| CSF-1 | P09603 |

**Table S9. Distribution of FlowSOM defined NK cell metaclusters in $IPI^-$ and $IPI^+$ patients**

| FlowSOM Metaclusters | SRN | | D7 | | D14 | | D28 | |
|---|---|---|---|---|---|---|---|---|
| | $IPI^-$ (n=6) (%) | $IPI^+$ (n=3) (%) | $IPI^-$ (n=6) (%) | $IPI^+$ (n=2) (%) | $IPI^-$ (n=6) (%) | $IPI^+$ (n=3) (%) | $IPI^-$ (n=4) (%) | $IPI^+$ (n=3) (%) |
| **cluster 1** | 15.0 | 28.8 | 18 | 27.2 | **1.48** | **19.6*** | **4.01** | **16.1** |
| **cluster 2** | 36.3 | 30.0 | 44.2 | 44.7 | **24.1** | **45.6** | **24.9** | **42.6** |
| **cluster 3** | 1.23 | 1.1 | 1.98 | 1.69 | 3.35 | 2.03 | 3.99 | 4.12 |
| **cluster 4** | 0.96 | 0.67 | 2.4 | 3.23 | **3.68** | **0.82** | 3.10 | 2.23 |
| **cluster 5** | 4.15 | 5.26 | 1.69 | 1.21 | 4.40 | 2.79 | 7.08 | 4.9 |
| **cluster 6** | 0.048 | 0.30 | 0.64 | 0.49 | 0.02 | 0.07 | 0.25 | 0.39 |
| **cluster 7** | 1.36 | 1.32 | 1.78 | 2.02 | 2.95 | 1.01 | 2.88 | 1.71 |
| **cluster 8** | 39.5 | 30.6 | 26.1 | 21.1 | **58.9** | **26.2** | **53** | **24** |
| **cluster 9** | 1.46 | 2.84 | 0.56 | 0.42 | 1.14 | 1.94 | 0.67 | 3.91 |
| **cluster 10** | 0.0 | 0.025 | 2.71 | 0 | 0 | 0 | 0 | 0.01 |

*Median percentage. Clusters correspond to those defined in Figure S10

SRN: trial screening time point, D7: day +7, D14: day +14, D28: day +28, Non Resp: non-responder, Resp: responder based on RECIST v1.1 criteria.

**Table S10. Downregulated gene expression of IPI vs no IPI on NK cells on day +7 after CIML NK infusion.**

| | baseMean | log2FoldChange | lfcSE | stat | p-value | padj |
|---|---|---|---|---|---|---|
| CADM2 | 55.04647 | -7.60108 | 1.576754 | -4.82072 | 1.43E-06 | 0.000525 |
| TMSB4Y | 37.05154 | -5.0083 | 1.033339 | -4.84671 | 1.26E-06 | 0.000474 |
| AC244213.1 | 21.2137 | -4.17627 | 1.204848 | -3.46622 | 0.000528 | 0.033996 |
| HLA-DRB5 | 6214.618 | -3.83062 | 1.032639 | -3.70954 | 0.000208 | 0.019036 |
| CEBPA | 53.83618 | -3.26463 | 0.968418 | -3.3711 | 0.000749 | 0.042486 |
| PINLYP | 112.5701 | -3.16717 | 0.839266 | -3.77373 | 0.000161 | 0.016418 |
| ARHGEF10 | 144.2845 | -2.98351 | 0.633346 | -4.71072 | 2.47E-06 | 0.000859 |
| LGR6 | 51.98837 | -2.73224 | 0.762366 | -3.5839 | 0.000339 | 0.026174 |
| AC068587.4 | 364.8396 | -2.5293 | 0.412908 | -6.12557 | 9.04E-10 | 1.50E-06 |
| AK5 | 175.7575 | -2.42999 | 0.556229 | -4.36868 | 1.25E-05 | 0.002467 |
| SLC1A7 | 78.29629 | -2.34158 | 0.67818 | -3.45274 | 0.000555 | 0.034939 |
| AL035701.1 | 141.5134 | -2.31104 | 0.417696 | -5.53284 | 3.15E-08 | 2.60E-05 |
| LINC02610 | 45.004 | -2.30797 | 0.667704 | -3.45657 | 0.000547 | 0.034778 |
| FCRL6 | 97.85369 | -2.29907 | 0.495849 | -4.63663 | 3.54E-06 | 0.001064 |
| ZMAT4 | 206.8177 | -2.29644 | 0.554648 | -4.14036 | 3.47E-05 | 0.005094 |
| PTGDS | 537.2453 | -2.09712 | 0.38677 | -5.42213 | 5.89E-08 | 3.89E-05 |
| AL355581.1 | 128.1178 | -1.9757 | 0.553205 | -3.57137 | 0.000355 | 0.027243 |
| GRAP | 68.3335 | -1.9637 | 0.576554 | -3.40593 | 0.000659 | 0.038886 |
| KIR3DL1 | 6401.976 | -1.86837 | 0.279607 | -6.68211 | 2.36E-11 | 7.79E-08 |
| JAKMIP2 | 457.1836 | -1.84826 | 0.268346 | -6.8876 | 5.67E-12 | 2.50E-08 |
| AC007038.1 | 101.3398 | -1.84793 | 0.53736 | -3.43891 | 0.000584 | 0.036426 |
| P2RY11 | 294.366 | -1.80654 | 0.40473 | -4.46357 | 8.06E-06 | 0.001838 |
| LIM2 | 683.0227 | -1.78475 | 0.254244 | -7.01984 | 2.22E-12 | 1.47E-08 |
| AC026347.1 | 103.8533 | -1.78135 | 0.513855 | -3.46663 | 0.000527 | 0.033996 |
| MMP23B | 227.8762 | -1.69653 | 0.487032 | -3.48341 | 0.000495 | 0.032894 |
| COLQ | 346.4287 | -1.68213 | 0.308991 | -5.44393 | 5.21E-08 | 3.83E-05 |
| PTCH1 | 160.9171 | -1.63751 | 0.488573 | -3.35161 | 0.000803 | 0.044722 |
| LINC01504 | 277.1627 | -1.62518 | 0.39372 | -4.12775 | 3.66E-05 | 0.005323 |
| DOK7 | 180.6451 | -1.61975 | 0.436109 | -3.7141 | 0.000204 | 0.018855 |
| DHX58 | 199.7273 | -1.6148 | 0.477517 | -3.38166 | 0.00072 | 0.041356 |
| CXCR2 | 151.52 | -1.61171 | 0.391388 | -4.11794 | 3.82E-05 | 0.005494 |
| KIR2DL1 | 1921.041 | -1.53236 | 0.275547 | -5.56116 | 2.68E-08 | 2.36E-05 |
| FGL2 | 222.9219 | -1.5238 | 0.370318 | -4.11485 | 3.87E-05 | 0.005508 |
| CCDC200 | 152.6728 | -1.50137 | 0.425912 | -3.52508 | 0.000423 | 0.029595 |
| FASLG | 382.6397 | -1.49589 | 0.397247 | -3.76564 | 0.000166 | 0.016767 |
| MYO15B | 181.4852 | -1.45669 | 0.350805 | -4.15244 | 3.29E-05 | 0.004887 |
| TNFAIP8L2 | 660.4027 | -1.45615 | 0.411806 | -3.536 | 0.000406 | 0.029424 |
| LAIR2 | 2956.429 | -1.45162 | 0.425993 | -3.40762 | 0.000655 | 0.038886 |
| RTP4 | 417.0125 | -1.40379 | 0.346049 | -4.05663 | 4.98E-05 | 0.006857 |
| APBA2 | 282.4338 | -1.39377 | 0.405855 | -3.43415 | 0.000594 | 0.036647 |

| | | | | | | |
|---|---|---|---|---|---|---|
| LINC02384 | 791.8301 | -1.37716 | 0.274436 | -5.01813 | 5.22E-07 | 0.000225 |
| GIMAP5 | 1510.298 | -1.32531 | 0.364196 | -3.63901 | 0.000274 | 0.023049 |
| KIF16B | 153.783 | -1.25358 | 0.376897 | -3.32606 | 0.000881 | 0.04873 |
| L3MBTL4 | 204.8273 | -1.22892 | 0.365668 | -3.36076 | 0.000777 | 0.043919 |
| SLC4A8 | 238.6469 | -1.19642 | 0.316948 | -3.77483 | 0.00016 | 0.016418 |
| PLEKHF1 | 1255.362 | -1.19472 | 0.297374 | -4.01756 | 5.88E-05 | 0.007775 |
| ALOX5AP | 1846.787 | -1.19429 | 0.355664 | -3.35792 | 0.000785 | 0.044185 |
| AC093010.2 | 273.2598 | -1.19337 | 0.31264 | -3.81706 | 0.000135 | 0.014637 |
| AC009093.2 | 683.7193 | -1.12289 | 0.332246 | -3.3797 | 0.000726 | 0.041356 |

**Table S11. Upregulated gene expression of IPI vs no IPI on NK cells on day +7 after CIML NK infusion.**

| | baseMean | log2FoldChange | lfcSE | stat | p-value | padj |
|---|---|---|---|---|---|---|
| IGLV2-8 | 144.2895 | 24.17025 | 4.785172 | 5.051072 | 4.39E-07 | 0.0002 |
| IGHA2 | 108.7636 | 23.76646 | 4.785311 | 4.966545 | 6.82E-07 | 0.000273 |
| IGLV7-46 | 100.0188 | 23.67232 | 4.78536 | 4.946821 | 7.54E-07 | 0.000293 |
| XIST | 1023.665 | 13.63114 | 1.496672 | 9.107631 | 8.42E-20 | 1.11E-15 |
| TSIX | 79.6045 | 7.511076 | 1.226371 | 6.124637 | 9.09E-10 | 1.50E-06 |
| CAVIN3 | 48.79488 | 5.784021 | 0.983352 | 5.881942 | 4.05E-09 | 5.36E-06 |
| TRGV5P | 17.75059 | 4.68782 | 1.249642 | 3.75133 | 0.000176 | 0.017228 |
| RAMACL | 54.74526 | 4.305536 | 1.194565 | 3.604272 | 0.000313 | 0.024636 |
| PPBP | 58.05265 | 4.171809 | 1.225171 | 3.405083 | 0.000661 | 0.038886 |
| PRSS2 | 63.78106 | 3.810247 | 1.098174 | 3.46962 | 0.000521 | 0.033996 |
| ADGRE2 | 61.44864 | 3.705851 | 0.828392 | 4.473548 | 7.69E-06 | 0.001838 |
| KIF26A | 21.98071 | 3.539905 | 0.95257 | 3.716161 | 0.000202 | 0.018834 |
| LRP5 | 33.13186 | 3.141966 | 0.871184 | 3.606546 | 0.00031 | 0.024568 |
| AC246817.2 | 30.20989 | 3.051681 | 0.813936 | 3.749287 | 0.000177 | 0.017241 |
| TOX2 | 39.73808 | 2.997282 | 0.816087 | 3.67275 | 0.00024 | 0.021293 |
| TMEM98 | 39.59011 | 2.895226 | 0.761206 | 3.803474 | 0.000143 | 0.015123 |
| CARD17 | 72.70379 | 2.799173 | 0.681905 | 4.104931 | 4.04E-05 | 0.005689 |
| AC246817.1 | 127.8617 | 2.592762 | 0.485484 | 5.340568 | 9.27E-08 | 5.83E-05 |
| SNCA | 145.7029 | 2.485349 | 0.555864 | 4.471144 | 7.78E-06 | 0.001838 |
| GPAT3 | 80.85746 | 2.284091 | 0.596985 | 3.826045 | 0.00013 | 0.014348 |
| APP | 68.11189 | 2.034386 | 0.570262 | 3.567461 | 0.00036 | 0.027243 |
| DOCK7 | 88.31597 | 1.950557 | 0.458809 | 4.251346 | 2.12E-05 | 0.003797 |
| SNHG17 | 1129.81 | 1.879422 | 0.362145 | 5.189701 | 2.11E-07 | 0.000103 |
| HOXA9 | 104.561 | 1.839826 | 0.488929 | 3.762971 | 0.000168 | 0.016819 |
| PYCR1 | 253.5039 | 1.820596 | 0.39003 | 4.667835 | 3.04E-06 | 0.000972 |
| YBX3 | 664.2052 | 1.793945 | 0.346538 | 5.176757 | 2.26E-07 | 0.000107 |
| KIR2DL4 | 3949.732 | 1.687955 | 0.363635 | 4.641893 | 3.45E-06 | 0.001062 |
| MZB1 | 748.4818 | 1.670881 | 0.305715 | 5.465478 | 4.62E-08 | 3.59E-05 |
| PPP1R9A | 513.896 | 1.539628 | 0.38195 | 4.030967 | 5.55E-05 | 0.007572 |
| BAG3 | 152.3445 | 1.503187 | 0.37356 | 4.023948 | 5.72E-05 | 0.007721 |
| IL32 | 25643.01 | 1.494879 | 0.343742 | 4.348845 | 1.37E-05 | 0.002585 |
| UCHL1 | 117.2306 | 1.473745 | 0.421822 | 3.49376 | 0.000476 | 0.032274 |
| CCNA2 | 1390.505 | 1.426105 | 0.21545 | 6.619201 | 3.61E-11 | 9.55E-08 |
| ANLN | 694.0548 | 1.407808 | 0.280643 | 5.016361 | 5.27E-07 | 0.000225 |
| PDIA5 | 176.129 | 1.393854 | 0.400051 | 3.484188 | 0.000494 | 0.032894 |
| TTK | 572.5735 | 1.393654 | 0.36165 | 3.853602 | 0.000116 | 0.013267 |
| PLK1 | 1469.016 | 1.387198 | 0.245975 | 5.639582 | 1.70E-08 | 1.65E-05 |
| POGLUT2 | 179.6014 | 1.338272 | 0.345736 | 3.870788 | 0.000108 | 0.012694 |

| | | | | | | |
|---|---|---|---|---|---|---|
| CDK2AP1 | 1116.68 | 1.31833 | 0.248906 | 5.296506 | 1.18E-07 | 6.50E-05 |
| KIF23 | 625.033 | 1.313937 | 0.250199 | 5.251568 | 1.51E-07 | 7.98E-05 |
| FBXO5 | 1309.244 | 1.312604 | 0.285058 | 4.604694 | 4.13E-06 | 0.001214 |
| SKA3 | 612.637 | 1.307643 | 0.278978 | 4.68726 | 2.77E-06 | 0.000939 |
| CXCR3 | 2488.348 | 1.300967 | 0.311895 | 4.171164 | 3.03E-05 | 0.004659 |
| MYBL2 | 1575.72 | 1.29519 | 0.220997 | 5.860678 | 4.61E-09 | 5.54E-06 |
| HOXA10 | 235.4822 | 1.280209 | 0.342779 | 3.734793 | 0.000188 | 0.017871 |
| HSPD1 | 6215.493 | 1.275441 | 0.200733 | 6.353909 | 2.10E-10 | 4.63E-07 |
| UBE2C | 3078.726 | 1.269929 | 0.239731 | 5.297317 | 1.18E-07 | 6.50E-05 |
| CDK1 | 1115.939 | 1.255663 | 0.222834 | 5.634979 | 1.75E-08 | 1.65E-05 |
| BIRC5 | 2087.584 | 1.247317 | 0.214617 | 5.811819 | 6.18E-09 | 6.81E-06 |

**Table S12. Panel of HLA antibodies for donor chimerism.**

| Antibody | Fluorochrome | Clone | Amount (uL) | Vendor | Catalog # |
|---|---|---|---|---|---|
| BW4 | FITC | REA274 | 5.0 | Miltenyi | 130-103-846 |
| BW6 | FITC | REA143 | 1.0 | Miltenyi | 130-123-264 |
| A2 | FITC | BB7.2 | 2.5 | Biolegend | 343304 |
| A3 | FITC | GAP.A3 | 2.5 | eBioscience | 501122136 |

| HLA | patient num | Target staining |
|---|---|---|
| A3 | 1 | Recipient |
| Bw6 | 2 | Recipient |
| none‡ | 3 | |
| A3 | 4 | donor |
| A2 | 5 | Recipient |
| A3 | 6 | donor |
| Bw4 | 7 | donor |
| Bw4 | 8 | donor |
| Bw6 | 9 | donor |
| *Bw4 | 10 | donor |

‡ No commercially available anti-HLA antibody that was expected to distinguish donor vs recipient

*HLA staining couldn't differentiate between donor and recipient cells

**Table S13. p-values for NK and T cell subset comparisons**

| | CD56$^+$ | CD56$^+$ Dim | CD56$^+$ Bright | CD56$^+$ dim NKG2A$^+$ | CD56$^+$ dim KIR$^+$ | CD56$^+$ dim CD57$^+$ | CD3$^+$ | CD4$^+$ Tcon | CD4$^+$ Tregs |
|---|---|---|---|---|---|---|---|---|---|
| SRN vs. 7 | <0.0001 | 0.6006 | 0.9997 | 0.9478 | 0.956 | 0.844 | 0.0059 | 0.1372 | <0.0001 |
| SRN vs. 14 | 0.0152 | 0.0302 | 0.023 | 0.9967 | >0.99 | 0.0117 | 0.4275 | 0.1628 | 0.0573 |
| SRN vs. 28 | 0.0664 | 0.2503 | 0.7208 | 0.9453 | >0.99 | 0.0673 | 0.9291 | 0.0515 | >0.99 |
| SRN vs. 42 | 0.2003 | 0.3701 | 0.8096 | 0.7687 | >0.99 | 0.0598 | 0.9919 | 0.8871 | >0.99 |
| SRN vs. D60 | 0.9384 | 0.0275 | 0.9985 | 0.9697 | 0.99 | 0.1019 | 0.9128 | 0.0918 | 0.9836 |

| | CD8$^+$ | Treg vs. Tcon | CD4$^+$ | CD4$^+$ Naïve | CD4$^+$ EM | CD4$^+$ CM | CD8$^+$ Naïve | CD8$^+$ EM | CD8$^+$ CM |
|---|---|---|---|---|---|---|---|---|---|
| SRN vs. 7 | 0.4019 | <0.0001 | 0.9282 | 0.0328 | 0.9968 | 0.2121 | 0.2454 | 0.9984 | 0.9955 |
| SRN vs. 14 | 0.6726 | 0.0022 | 0.7462 | 0.0028 | 0.862 | 0.1561 | 0.1046 | 0.7492 | 0.019 |
| SRN vs. 28 | 0.0082 | 0.9941 | 0.0587 | 0.0015 | 0.4938 | 0.0465 | 0.7694 | 0.2712 | 0.0005 |
| SRN vs. 42 | 0.0749 | 0.9727 | 0.4673 | 0.0041 | 0.8076 | 0.7238 | 0.9735 | 0.3031 | 0.0416 |
| SRN vs. D60 | 0.0044 | 0.9997 | 0.1129 | 0.0157 | 0.6633 | 0.1557 | 0.9721 | 0.07 | 0.0029 |

*adjusted p-values are determined using Dunnett's multiple comparisons test

**Table S14. p-values for dot plots figures**

| P-values for Figure 6D | D7 $IPI^{+}$ vs $IPI^{-}$ | D28 $IPI^{+}$ vs $IPI^{-}$ |
|---|---|---|
| PDCD1 | 9.6E-01 | 1.0E+00 |
| CTLA4 | 8.7E-01 | 8.8E-01 |
| TIGIT | 7.8E-03 | 1.0E+00 |
| HAVCR2 | 5.2E-04 | 1.0E+00 |
| TOX | 4.2E-01 | 9.9E-01 |
| LAG3 | 1.6E-03 | 9.9E-01 |
| ENTPD1 | 3.5E-02 | 8.0E-01 |
| TCF7 | 9.8E-01 | 1.6E-05 |
| IL7R | 1.0E+00 | 8.5E-07 |
| SELL | 2.7E-01 | 6.8E-02 |
| CCR7 | 9.3E-01 | 7.6E-02 |
| CD28 | 9.1E-01 | 2.8E-01 |
| HLA-E | 1.0E+00 | 1.1E-01 |
| LGALS9 | 6.1E-01 | 9.9E-01 |
| PVR | 2.7E-01 | 8.8E-01 |
| CD274 | 2.4E-01 | 4.0E-01 |
| PDCD1LG2 | 1.0E+00 | 5.4E-01 |
| S100A8 | 5.9E-01 | 3.5E-04 |
| S100A9 | 4.8E-01 | 2.3E-04 |
| CLEC2D | 1.7E-01 | 1.4E-01 |
| MICA | 6.4E-01 | 2.3E-01 |
| MICB | 9.4E-01 | 6.0E-01 |
| ULBP1 | 4.7E-01 | 4.5E-01 |
| NCR3LG1 | 3.4E-01 | 3.0E-01 |
| HLA-G | 9.6E-08 | 6.2E-18 |
| ICAM1 | 4.0E-01 | 3.8E-02 |
| CD48 | 6.3E-03 | 3.4E-10 |

| **P-values for Supplementary Figure 3** | **Day 7 vs. D28** | |
|---|---|---|
| IL7R | 3.9E-99 | |
| SELL | 2.6E-25 | |
| KLRC1 | 2.7E-01 | |
| GZMK | 0.0E+00 | |
| CD44 | 1.1E-95 | |
| XCL1 | 6.5E-23 | |
| XCL2 | 1.7E-16 | |
| FCGR3A | 0.0E+00 | |
| CD160 | 1.1E-154 | |
| NCAM1 | 2.4E-23 | |
| KLRC2 | 2.0E-33 | |
| CD52 | 2.1E-64 | |
| CD3E | 2.6E-196 | |
| CD3D | 8.0E-01 | |
| CD3G | 5.1E-01 | |
| IL32 | 8.2E-06 | |
| FCER1G | 7.1E-242 | |
| ZEB2 | 0.0E+00 | |
| TBX21 | 3.9E-242 | |
| GZMB | 0.0E+00 | |
| PRF1 | 0.0E+00 | |
| NCR3 | 2.6E-64 | |
| KIR3DL1 | 2.6E-102 | |
| **P-values for Supplementary Figure 11** | **D7 $IPI^+$ vs $IPI^-$** | **D28 $IPI^+$ vs $IPI^-$** |
| JUN | 1.0E+00 | 1.0E+00 |
| JUNB | 2.1E-04 | 6.3E-14 |

| | | |
|---|---|---|
| FOS | 1.4E-01 | 2.3E-62 |
| FOSB | 6.5E-01 | 1.0E+00 |
| GZMB | 7.0E-140 | 1.0E+00 |
| PRF1 | 1.0E+00 | 1.0E+00 |
| MKI67 | 4.3E-124 | 7.8E-01 |
| CD274 | 2.2E-07 | 3.2E-01 |
| HAVCR2 | 1.6E-29 | 1.0E+00 |
| TIGIT | 1.0E+00 | 1.0E+00 |
| LAG3 | 5.7E-58 | 1.0E+00 |
| PDCD1 | 2.9E-01 | 1.8E-01 |
| CD226 | 1.0E+00 | 2.3E-01 |
| KLRK1 | 7.5E-11 | 4.1E-11 |
| NCR2 | 7.5E-02 | 1.0E+00 |
| KIR2DL4 | 0.0E+00 | 2.1E-15 |
| ITGAL | 1.0E+00 | 1.0E+00 |
| CD244 | 8.0E-01 | 1.0E+00 |
| CD80 | 2.1E-10 | 3.4E-01 |
| KLRC1 | 5.8E-40 | 1.3E-33 |
| KLRD1 | 1.0E+00 | 1.0E+00 |
| KIR3DL1 | 1.0E+00 | 1.0E+00 |
| KIR3DL2 | 4.1E-01 | 1.0E+00 |
| KIR3DL3 | 1.2E-09 | 4.0E-01 |
| CEACAM1 | 3.7E-02 | 2.8E-01 |
| FAS | 1.0E-16 | 6.6E-02 |
| TIMD4 | 4.8E-11 | 9.7E-01 |
| LILRB1 | 1.0E+00 | 9.9E-01 |
| LILRB5 | 7.9E-01 | 6.5E-01 |
| KLRG1 | 1.0E+00 | 1.0E+00 |
| KLRB1 | 1.0E+00 | 1.0E+00 |

**Table S15. p-values for Figure 6E**

| | $CD8^+$ T Cells | $CD8^+$ T Cells-like | DN T Cells | Tcons | Tregs |
|---|---|---|---|---|---|
| TIGIT | 1.2E-13 | 2.0E-11 | 8.1E-01 | 1.8E-01 | 4.0E-04 |
| LAG3 | 4.7E-02 | 4.3E-09 | 5.5E-09 | 2.1E-01 | 4.9E-02 |
| HAVCR2 | 6.5E-42 | 1.6E-19 | 2.8E-05 | 4.1E-01 | 1.3E-02 |

*all comparisons are between Day +7 (D7) and Day +28 (D28)

**Table S16. p-values for Supplementary Figure 16**

| | CCL3 D7 | CCL3 D28 | CCL4 D7 | CCL4 D28 |
|---|---|---|---|---|
| $NK.CD16^+$ | 4.7E-02 | 1.3E-05 | 8.1E-01 | 8.7E-20 |
| $NK.CD16^-$ | 8.2E-01 | 7.6E-12 | 9.7E-02 | 1.3E-48 |
| $CD8^+$ T Cells | 2.9E-01 | 1.6E-05 | 3.1E-02 | 1.1E-02 |
| $CD8^+$ T Cells-like | 7.0E-01 | 5.2E-01 | 4.6E-01 | 7.2E-01 |
| DN T Cells | 7.0E-01 | 1.3E-02 | 3.0E-01 | 1.7E-02 |
| Platelets | NA | 8.2E-01 | NA | 7.1E-01 |
| Tregs | 8.4E-01 | 7.8E-01 | 6.0E-01 | 6.0E-01 |
| Tcons | 1.0E+00 | 8.7E-01 | 7.9E-01 | 8.8E-01 |
| Classical Monocytes | 6.9E-01 | 3.8E-05 | 5.2E-01 | 2.5E-43 |
| Non-Classical Monocytes | NA | 2.5E-01 | NA | 1.9E-01 |
| B Cells | NA | 8.9E-01 | NA | 5.9E-01 |

*all comparisons are between $IPI^+$ and $IPI^-$ conditions

**Table S17. Representativeness of Study Participants**

| Cancer type | Relapsed/refractory head and neck cancer |
|---|---|
| Considerations related to: | |
| Sex | Head and neck carcinoma (HNC) is the sixth most common cancer worldwide, with approximately 70,000 new cases reported in the US annually, accounting for 3% of all malignancies. The incidence is much higher in men (17.2 per 100,000) compared to women, with an estimated ratio ranging from 2:1 – 4:1. This higher incidence parallels the relative incidence of modifiable behaviors between the sexes that are known risk factors of HNC, including smoking and alcohol use. |
| Age | HNC can be associated with viruses such as HPV and EBV, and the median age at diagnosis differs between virus-associated and unassociated cases. Among non-virally associated cases, the median age at diagnosis is approximately 66 years, while in HPV and EBV-associated cases, the median age at diagnosis is around 50-53 years. |
| Race/ethnicity | In the USA, the overall incidence of HNC is higher among the African American population as compared to others, with the latter particularly affected by laryngeal cancers. HNC mortality is significantly higher among African American men. |
| Geography | World-wide, the geographic distribution of HNC reflects a combination of genetic and behavioral risk factors associated with the disease. In the USA, the geographic distribution trends with use of tobacco and alcohol consumption, with heavy users of both substances having a greater than 30-fold risk of developing HNC. |
| Other considerations | While the mortality due to HNC has fallen considerably over the last 50 years in the US, significant disparity in survival exist depending on the stage of diagnosis. Earlier detection due to screening in communities with more access to these programs results in improved survival, as compared to those communities (for example, with low socioeconomic status or rural) who do not have adequate access to such programs. |
| Overall representativeness of this study | The age distribution of our study is representative of a population of relapsed/refractory HNC. Median age was 58, which agrees with the predominance of virally associated HNC (8/10 patients were HPV+) and their earlier age at diagnosis. Nine of the 10 treated patients |

| | |
|---|---|
| | were men, consistent with male predominance in this disease. Only 1/10 treated patients was African American, deviating from the expected overall incidence of this disease. This could be explained by the majority of our patients having oropharyngeal primaries (as opposed to laryngeal), the result of a small sample size in phase 1, as well as the study population being at one treatment site (Dana-Farber Cancer Institute in Boston, MA). |
